# Disintegration Fingerprinting: A Low-Cost and User-Friendly Tool for Identifying Substandard and Falsified Solid-Dosage Medicines

**DOI:** 10.1101/2025.08.15.25333621

**Authors:** Ishmam Fatima, Oscar Fajardo, Canhui Liu, Harshith Sadhu, William H. Grover

## Abstract

Substandard and falsified medicines cause thousands of deaths and waste billions of dollars worldwide every year. There is an urgent need for low-cost and simple-to-use tools for identifying these “fake drugs.” In this work we demonstrate “Disintegration Fingerprinting” (DF), a technique that identifies pills, tablets, caplets, and other solid-dosage drugs based on how the drug disintegrates and dissolves in liquid. The DF hardware consists of a water-filled transparent plastic cup atop a conventional magnetic stirrer. An inexpensive sensor mounted on the outside of the cup shines infrared light into the cup and measures the amount of light that is reflected back to the sensor. When a pill is added to the stirred water, the pill begins to disintegrate into particles that swirl around inside the cup. Whenever one of these particles passes near the infrared sensor, the particle reflects additional light back to the sensor and creates a millisecond-duration peak in a plot of sensor output versus time. The number of particles in the water changes over time as the particles continue to disintegrate and (for some samples) eventually dissolve away. By plotting the number of particles detected versus time, we create a Disintegration Fingerprint that can be used to identify the drug product. In a proof-of-concept study, we used DF to analyze pills from 32 different drug products (including antibiotics, opioid and non-opioid analgesics, antidepressants, anti-inflammatories, antiemetics, antihistamines, decongestants, muscle relaxants, expectorants, sleep aids, cold medicines, antacids, hormonal birth control, and dietary supplements, as well as a simulated falsified drug product). We found that DF correctly identified 90% of these pills, and the technique can even distinguish name-brand and generic versions of the same drug. Even when testing pills from 33 different manufacturing lots of two similar drug products, purchased in eight different states or provinces in two countries, with manufacturing dates that span almost three years, some of which were intentionally subjected to extreme temperatures (50 °C or −20°C for 35 days), our technique still correctly identified 100% of the pills. By providing a fast (60-minute), inexpensive ($33 USD), and easy-to-use tool for identifying substandard and falsified medicines, Disintegration Fingerprinting can play an important role in the fight against fake drugs.

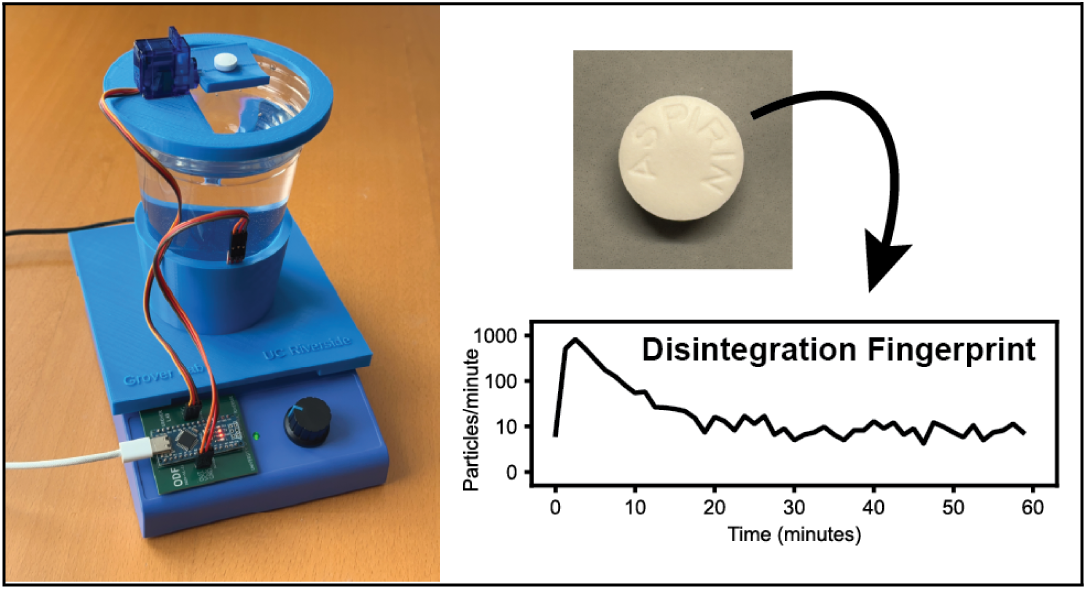

## Introduction

The World Health Organization (WHO) estimates that 1 in 10 medical products in low-and middle-income countries is substandard or falsified,^1^ meaning that the products fail to meet either their quality standards or their specifications, or both. Substandard or falsified (SF) medications often lack active pharmaceutical ingredients or contain incorrect dosages, making them ineffective or harmful. The global impact of SF medicines is enormous. An estimated 122,000 deaths of children under five years of age were linked to poor-quality antimalarial drugs in 2013.^2^ In low- and middle-income countries (LMICs), 19.1% of antimalarials and 12.4% of antibiotics were found to be substandard or falsified,^3^ and up to 30% of medicines in parts of Africa, Asia, and Latin America are counterfeit.^4^ SF medicines also have a significant economic impact, with an estimated $30 billion USD spent on SF medicines in LMICs annually.^1^

Tools for identifying SF medicines can play a crucial role in combating this problem. Several existing techniques for detecting SF medicines are summarized in Table 1. Some of these tools perform *chemical* analyses to detect the presence or absence of active ingredients. For example, techniques such as high-performance liquid chromatography (HPLC) and mass spectrometry (MS) are gold standards for identifying a wide variety of SF medicines.^5^ Noninvasive spectroscopic techniques such as Raman spectroscopy can even detect active ingredients through a drug’s unopened packaging.^6^ However, tools like HPLC, MS, and Raman require expensive equipment and specialized operator training; this limits their accessibility in resource-constrained settings.^4^ Low-cost techniques for detecting active ingredients have also been developed. For example, paper analytical devices (PADs)^7^ and microfluidic paper-based assays (μPADs)^8^ use chromatography and color changes to detect active ingredients, the “PharmaChk” instrument uses aptamers to identify active ingredients,^9,10^ and the “GPHF-Minilab” kit uses thin-layer chromatography to detect 125 different active ingredients.^11,12^ Tests like these are relatively low-cost and easy-to-use, but they require a supply of consumables (disposable devices, reagents, reference standards, etc.), must be tailored for targeting specific chemical compounds, and may not be applicable to all active ingredients.

**Table 1.** Summary of several existing techniques for identifying substandard and falsified medicines.

| **Technique** | **Hardware** | **Consumables** | **Ease of use** | **Ref** |
| --- | --- | --- | --- | --- |
| Disintegration test | **Low** (stopwatch and cup) | None | **Easy** (visual inspection and stopwatch use) | 29 |
| Thin-layer chromatography (TLC) kit | **Moderate** (TLC plates, solvent containers and portable kit) | TLC plates, reagents, solvents | **Moderate** (basic lab skills) | 29 |
| Low-cost capillary electrophoresis (CE) device | **Moderate** (custom CE device, lab power supply) | Buffers, capillaries | **Moderate** (training in analytical chemistry) | 30 |
| Colorimetry of tablets and packaging | **Low** (handheld colorimeter) | None | **Easy** (visual comparisons of color readings) | 31 |
| Paper test cards (PADs) | **Low** (paper cards) | Paper cards, water | **Easy** (visual or photo to interpret colors) | 32 |
| FDA CD3+ multispectral scanner | **Low** (handheld scanner with LED lights) | None | **Easy** (point-and-shoot operation) | 33 |
| Handheld Raman scanner | **High** (handheld Raman spectrometer) | None | **Easy** (built-in library yields rapid ID) | 33 |
| Raman + X-Ray CT imaging | **High** (Raman spectrometer + X-ray CT machine) | None | **High** (training in both spectrometer and CT X-ray) | 34 |
| BARDS acoustic dissolution | **Moderate** (acoustic analyzer with microphone and software) | Mild acid solvent (e.g. HCl) | **Easy** (~5 min test; no complex sample prep) | 18 |
| Polymer analysis of packaging | **High** (FTIR and differential scanning calorimeter instruments) | None | **Moderate** (training in spectroscopy and calorimetry) | 35 |
| Microwave cavity perturbation | **Moderate** (microwave RF analyzer and 3D-printed holders) | None | **Moderate** (experimental prototype, basic RF/electronics knowledge) | 36 |
| PowDew smartphone droplet analysis | **Low** (smartphone with camera and app) | A drop of water | **Easy** (smartphone app infers authenticity from droplet behavior) | 37 |
| PharmaChk aptamer identification | **Moderate** (handheld system) | Probe reagents, disposable cartridge | **Easy** (basic user training on field-ready device) | 10 |
| WHO checklist for visual inspection | **Low** (no hardware) | None | **Easy** (follow a checklist) | 13 |
| High-performance liquid chromatography (HPLC) | **High** (bench top HPLC system) | Organic solvents, reagents, columns | **Difficult** (experience in sample prep and HPLC) | 38 |
| Fourier transform infrared spectroscopy (FTIR) | **High** (IR spectrometer) | None | **Easy** (point-and-shoot scanning) | 39 |
| Near-infrared spectroscopy (NIR) | **High** (NIR spectrometer) | None | **Easy** (handheld analyzer) | 40 |
| Fluorescence | **High** (fluorimeter) | None | **Moderate** (training in spectroscopy) | 41 |
| Nuclear magnetic resonance spectroscopy (NMR) | **High** (NMR spectrometer) | Deuterated solvents; cryogens (liquid N<sub>2</sub>/He for magnet) | **Difficult** (training in NMR) | 42 |
| X-Ray diffraction (XRD) | **High** (XRD apparatus) | None | **Moderate** (training in XRD, experience interpreting results) | 43 |
| Mass spectrometry (MS) | **High** (MS instrument) | Solvents, carrier gases, standards | **Difficult** (training in MS, experience interpreting results) | 44 |
| Desorption electrospray ionization (DESI MS) | **High** (MS instrument) | Solvent for spray; nebulizing gas | **Difficult** (training in MS) | 45 |
| Direct analysis in real time (DART MS) | **High** (MS instrument) | Inert gas (e.g. He) | **Difficult** (training in MS) | 46 |
| Disintegration Fingerprinting (DF; this work) | **Low** (\$33-\$42 USD) | None | **Easy** (analysis is fully automated; results can be pass-fail or quantitative) | |

Other tools perform *physical* analyses to identify SF medicines. These techniques do not directly detect the active ingredient, but they have the advantage of being applicable to virtually all medicines and can be low-cost as well. The most basic form of physical analysis is visual inspection: examining a tablet’s markings, color, surface finish, packaging, and so on. WHO’s “Tool for Visual Inspection of Medicines” is a checklist that guides the user through a 35-step qualitative visual inspection process.^13^ For more *quantitative* physical data, the GPHF-Minilab includes calipers and a digital balance for measuring pill size and mass,^11^ handheld glossmeters can be used to measure pill surface roughness,^14^ smartphones can be used to detect inconsistencies indicative of poor manufacturing or deliberate obfuscation,^15^ and dried droplets of liquid medicines make unique patterns that can be identified using machine learning.^16^

Physical measurements of how a pill disintegrates and dissolves in liquid are particularly interesting because they can be influenced by both the chemical composition of the pill (for example, two pills with different ingredients may dissolve at different speeds, or disintegrate into different-sized particles) and the process used to manufacture the pill (for example, two pills manufactured on different pill presses may disintegrate at different rates). In perhaps the simplest method of this type, a pill is placed into a cup filled with water and a stopwatch is used to record how much time it takes for the pill to disintegrate. This method is included in the GPHF-MiniLab,^11^ and while it is extremely simple and low-cost, it provides only a single number (a disintegration/dissolution time) and thus has limited ability to distinguish different drug products. Other techniques seek to extract richer data from the disintegration and dissolution process. For example, the dissolution apparatuses described by the US Pharmacopeia are routinely used to measure the release of active ingredients over time as a pill disintegrates and dissolves;^17^ a plot of the amount of active ingredient released vs. time could be used to identify a drug product. Other techniques use sound recordings^18^ and imaging along with machine learning^19^ to monitor the dissolution process. However, while methods like these produce data-rich “fingerprints” from the pill disintegration/dissolution process, the cost of the required hardware makes them less suitable for use in resource-limited settings.

In this work we introduce “Disintegration Fingerprinting” (DF), a low-cost and user-friendly technique for generating unique digital “fingerprints” based on how a pill disintegrates and dissolves. DF counts the number of drug particles present in solution over time as a drug product disintegrates and dissolves. Instead of using costlier conventional approaches like Coulter counting or dynamic light scattering to count particles, our prototype DF instrument uses a $3.50 USD light sensor originally marketed for use in line-following toy robots. As a typical pill dissolves, the number of particles detected by our sensor initially rises (as the pill disintegrates into particles). Depending on the pill, the particle count may continue to rise (if the particles fragment into smaller particles), or drop (if the particles dissolve away), or remain unchanged (if the particles are insoluble). We call these plots of particle count versus time “Disintegration Fingerprints” because their shapes vary significantly across different drug products. Consequently, Disintegration Fingerprinting can be used to determine whether two pills are likely the same (if they have similar DFs) or are definitely different (if they have significantly different DFs). Moreover, by constructing a library of DFs for different drug products, Disintegration Fingerprinting could be used to identify a pill (and flag suspect pills for additional scrutiny) in customs offices, clinics, pharmacies, and other settings.

## Materials and Methods

### Hardware

The hardware needed for Disintegration Fingerprinting can vary depending on a user’s needs and available resources. At minimum, the technique requires a stirred water-filled vessel to contain the disintegrating medication, a sensor for monitoring the pill disintegration/dissolution process, and electronic hardware for recording the sensor output and calculating the DF.

Our prototype Disintegration Fingerprinting instrument (Figure 1) uses an optical sensor marketed for use in line-following toy robots (RedBot line follower sensor QRE1113GR; SparkFun Electronics, Boulder, CO; $4.15 USD). The sensor consists of an LED that emits infrared light (940 nm) placed beside a phototransistor; when an object is placed in front of the sensor, light from the LED is reflected or scattered by the object and detected by the phototransistor. The voltage output of the sensor is measured using an Arduino Nano microcontroller clone. The optical sensor is held against the side of a disposable transparent PET plastic cup (18 ounces; Solo Cup Company, Lake Forest, IL) with a height of 120 mm, top diameter of 100 mm, and base diameter of 64 mm. The cup is placed on a low-cost magnetic stirrer (INTLLAB, Greenwood, IN; $23.99 USD) and a 25.4 × 8 mm Teflon-coated magnetic stir bar is added. In total, this required hardware costs about $33 USD.

**Figure 1:**
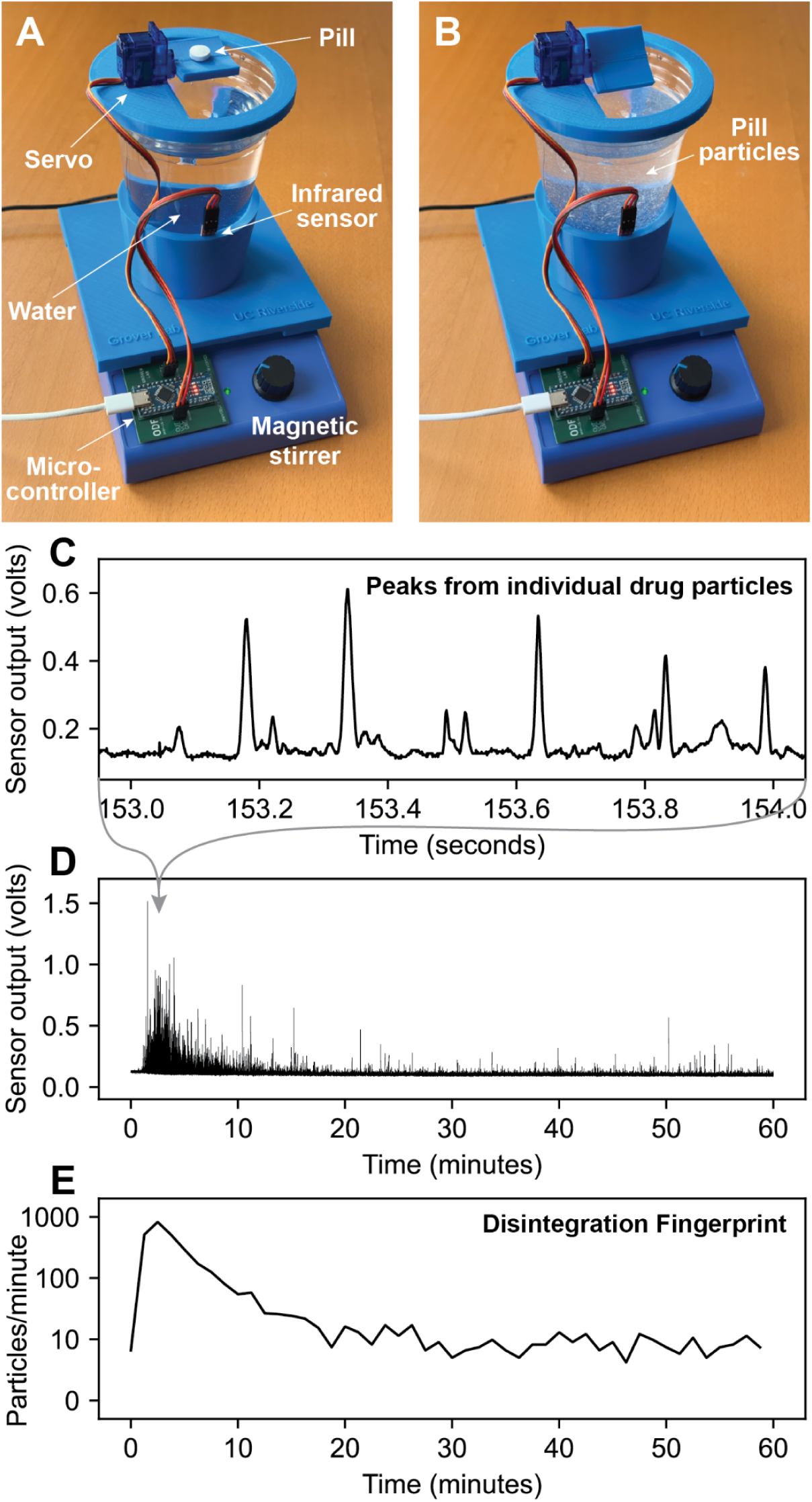
Our prototype Disintegration Fingerprinting apparatus. (**A**) A servo drops a single pill into a water-filled cup atop a magnetic stirrer. The pill begins to disintegrate into particles (**B**), and an infrared optical sensor on the side of the cup detects the light reflected by individual particles as they pass the sensor (**C**). As the particles disintegrate and dissolve further, the number of particles detected changes (**D**). A plot of peak count versus time (**E**) serves as a “Disintegration Fingerprint” for this pill.

For convenience and reproducibility, we also designed some optional custom components for the DF instrument. 3D-printed fixtures (shown in blue in Figure 1) hold the cup in place on the stirrer, hold the optical sensor at a fixed location against the side of the cup (3.5 cm from the bottom of the cup), and hold a servo above the cup. The servo (SG90; TowerPro Ltd, Taipei, Taiwan) automatically drops a pill into the cup at the desired time during a run. The 3D-printed fixtures were printed using a low-cost hobbyist-grade 3D printer (Ender 3, Creality, Shenzhen, China). Finally, a custom printed circuit board (shown in green in Figure 1) simplifies the electrical connections between the Arduino Nano clone, the optical sensor, and the pill-dropping servo. CAD files for each of these custom components are available for download.^20^ Adding this optional hardware increases the overall cost of our prototype DF instrument in Figure 1 to about $42 USD.

### Software

Custom software was used to program the Arduino Nano microcontroller clone; the software relays sensor measurements to the attached computer via USB and operates the pill-dropping servo at the desired time. Additional software (written in Python and running on the attached computer) records the sensor measurements, converts these measurements into Disintegration Fingerprints, performs comparisons between pairs of DFs, and generates the plots shown in this work. This software runs on both Windows and MacOS computers and is available for download.^20^

Our code for converting raw sensor voltage data into Disintegration Fingerprints follows several steps:

1. Very rarely, the computer may record individual voltage measurements from the Arduino Nano that are outside of the range of the microcontroller’s analog-to-digital converter (0 to 1023 arbitrary units, corresponding to 0 to 5 volts). This can be caused by electromagnetic interference from *e.g*. a nearby static electricity spark corrupting data on the interface between the microcontroller and the computer. We replaced these corrupted measurements with the previous valid measurement. In the 96-pill dataset analyzed in Figure 6, there are a total of about 8 × 10^8^ individual voltage measurements; only 45 of these measurements had to be replaced due to out-of-dynamic-range values, or about 0.000001% of the measurements.
2. The raw data was inverted (all measurements subtracted from 1023) to make particle peaks point in a positive (upward) direction.
3. Data before the 5-minute mark was discarded (only the data after the pill is added is analyzed).
4. Noise peaks are rare single voltage measurements that are significantly different from their neighboring measurements; they can be caused by spurious electrical signals like a static spark in the vicinity of the instrument or electrical noise created by the servo during motion. These noise peaks were detected using the SciPy^21^ function scipy.signal.find_peaks with a threshold of 20 (this identifies individual measurements that are more than 20 arbitrary units different from their immediately preceding and following measurements). The noise peak values were then removed and replaced by the average of the two neighboring measurements. Out of the approximately 8 × 10^8^ individual voltage measurements in the 96-pill dataset in Figure 6, only 136 measurements had to be replaced due to noise, or about 0.00002% of the measurements.
5. A moving linear regression of the sensor data was calculated to find baseline shifts indicative of bubble formation on the cup wall near the optical sensor. Bubbles are characterized by a slow rise in the baseline signal (as the bubble forms near the sensor) followed by a sudden drop in the baseline signal (as the bubble detaches and leaves the sensor area). To detect these bubble-induced baseline drops, we used a moving window with a width of 50 seconds. If the slope of the data in this moving window is ever less than −0.25 arbitrary units/second, then the run is flagged as containing a bubble and is discarded. In the 96-pill dataset in Figure 6, two of the runs were discarded due to bubble detection (one Microgestin Fe green pill, and one doxycycline pill) and replaced by analyzing another pill of the same product (these were both successful).
6. To locate peaks corresponding to drug particles passing the sensor, the scipy.signal.find_peaks function was used with a prominence threshold of 10.
7. To count drug particle peaks over time, the peak data was partitioned into bins of equal time lengths, then the number of peaks in each bin was divided by the bin duration to obtain the particles-per-second for that bin. The chosen bin duration determines the number of points in the resulting Disintegration Fingerprint; the bin duration used in this work (75 s) produces a DF with 48 points. Each bin’s peak count was increased by 1 (to avoid having values of 0 on a logarithmic plot) and plotted on a logarithmic axis vs. time to create the Disintegration Fingerprints shown in this work.

To calculate the similarity of two Disintegration Fingerprints, we subtracted the peak counts for each bin on the DFs, calculated the absolute value of these differences, and calculated the sum of the absolute values to create a single number, a “difference score.” These scores range from single digits (for two very similar DFs) to over 40,000 (for two very different DFs).

To find matches, we compared each pill’s Disintegration Fingerprint to every other pill’s DF in a given data set. For example, for the 96-pill dataset, each pill’s DF was compared to the other 95 pills (pills were not compared to themselves) and 95 difference scores were calculated. These comparisons were then ranked from lowest difference score (most similar) to highest (least similar). If a pill’s closest match was another pill of the same product, then we considered the pill successfully identified.

### Sample testing

Since purified laboratory water may not be readily available in all locations where Disintegration Fingerprinting might be used, all samples were analyzed in ordinary tap water from two locations in Riverside, California. Preliminary testing of DF with insoluble and inert particles was performed using 50 mg of red polyethylene microspheres (425-500 µm diameter, 1.0648 g/mL density; Cospheric, Santa Barbara, CA).

Approximately 100 µL of detergent (Palmolive dish soap; Colgate-Palmolive, New York, NY) was added to the water to keep the microspheres from clumping. The drug products tested were obtained in eight different states or provinces in two countries; their manufacturers and ingredients are shown in Table 2. The simulated falsified pills tested alongside the authentic drug products in Figure 6 were made by compressing powdered milk (Carnation instant nonfat dry milk, Nestle, Arlington, VA) using a 10-mm-diameter manual pill press (MUHWA Scientific, Shanghai, China).

**Table 2.**
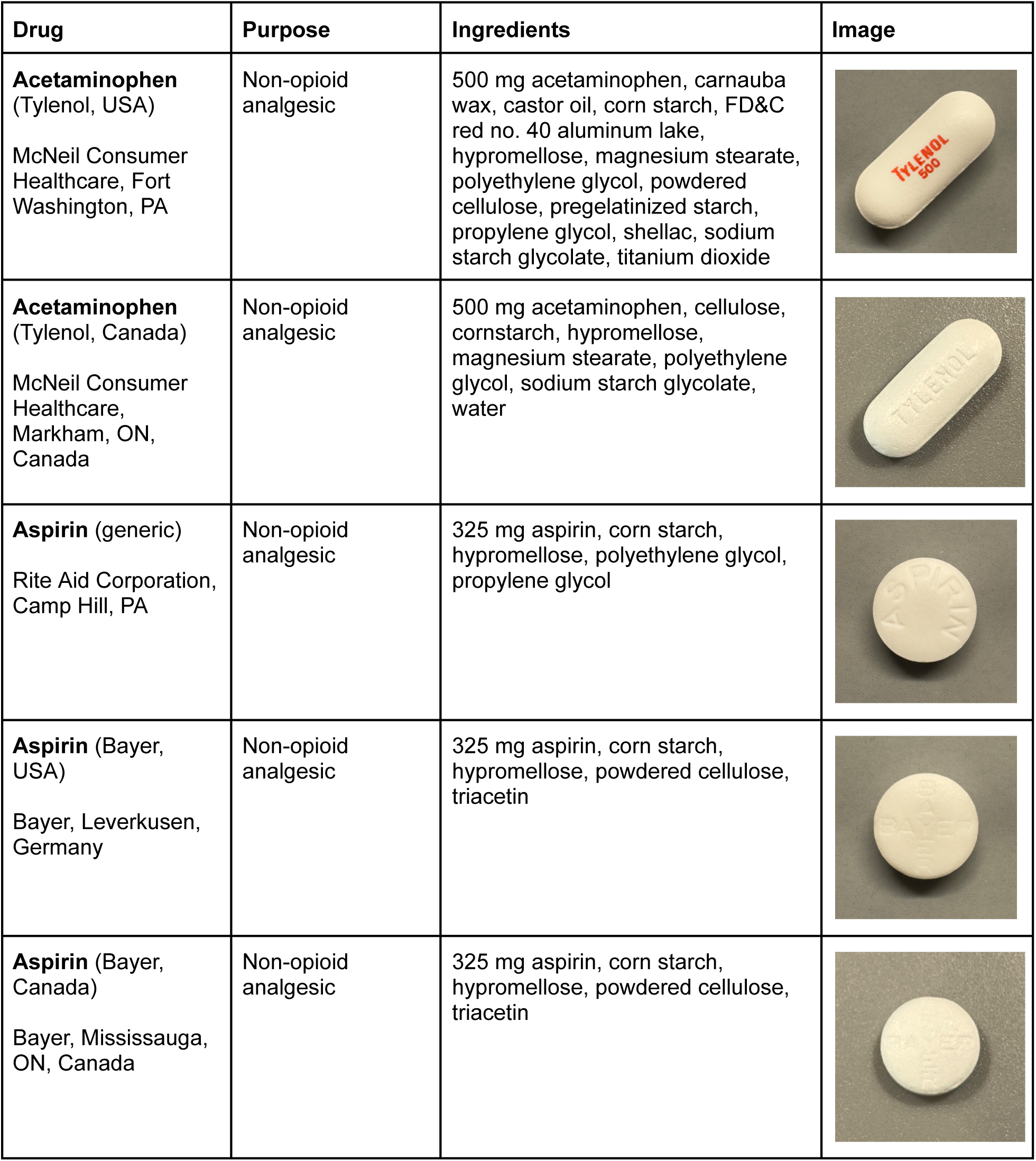
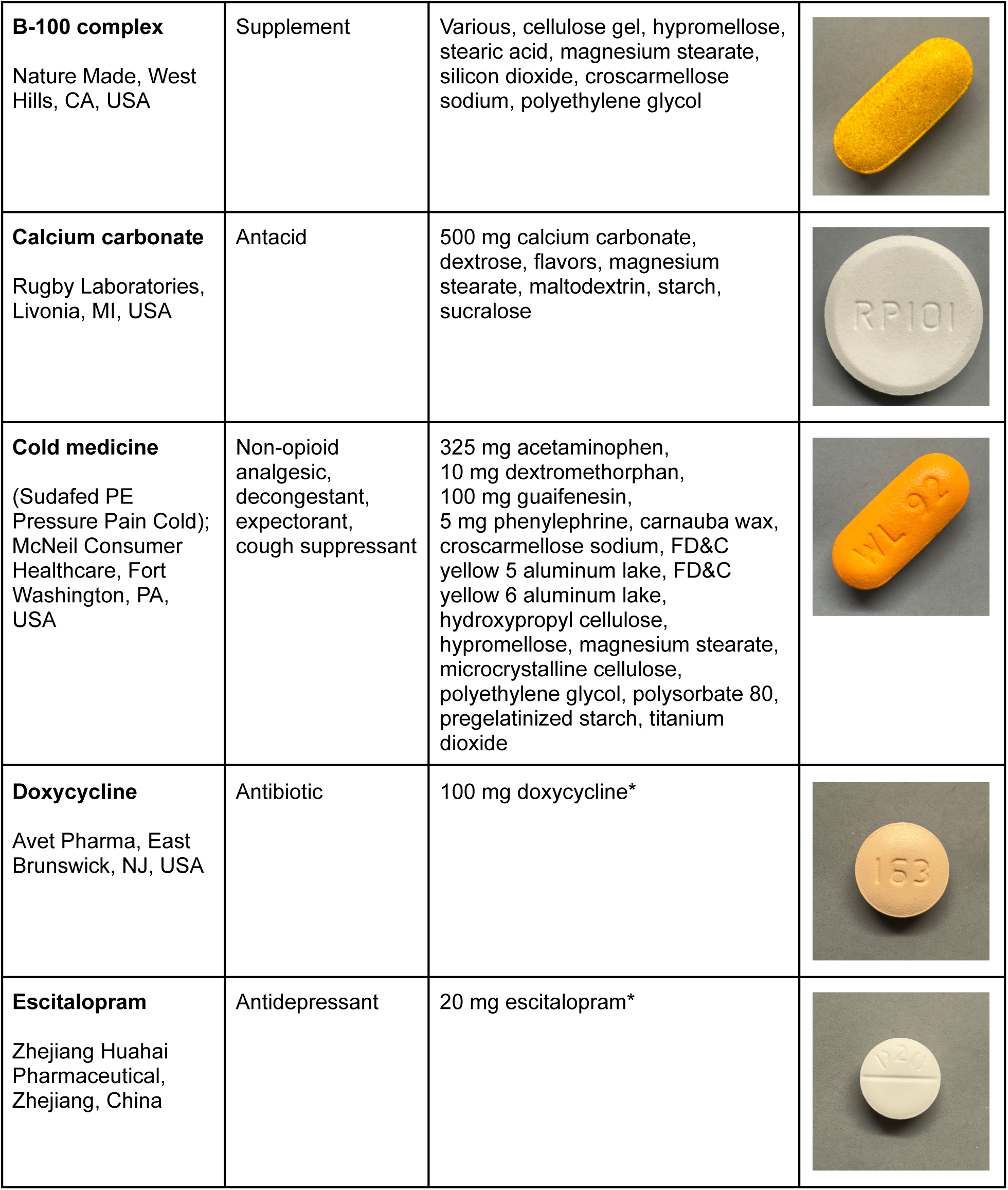
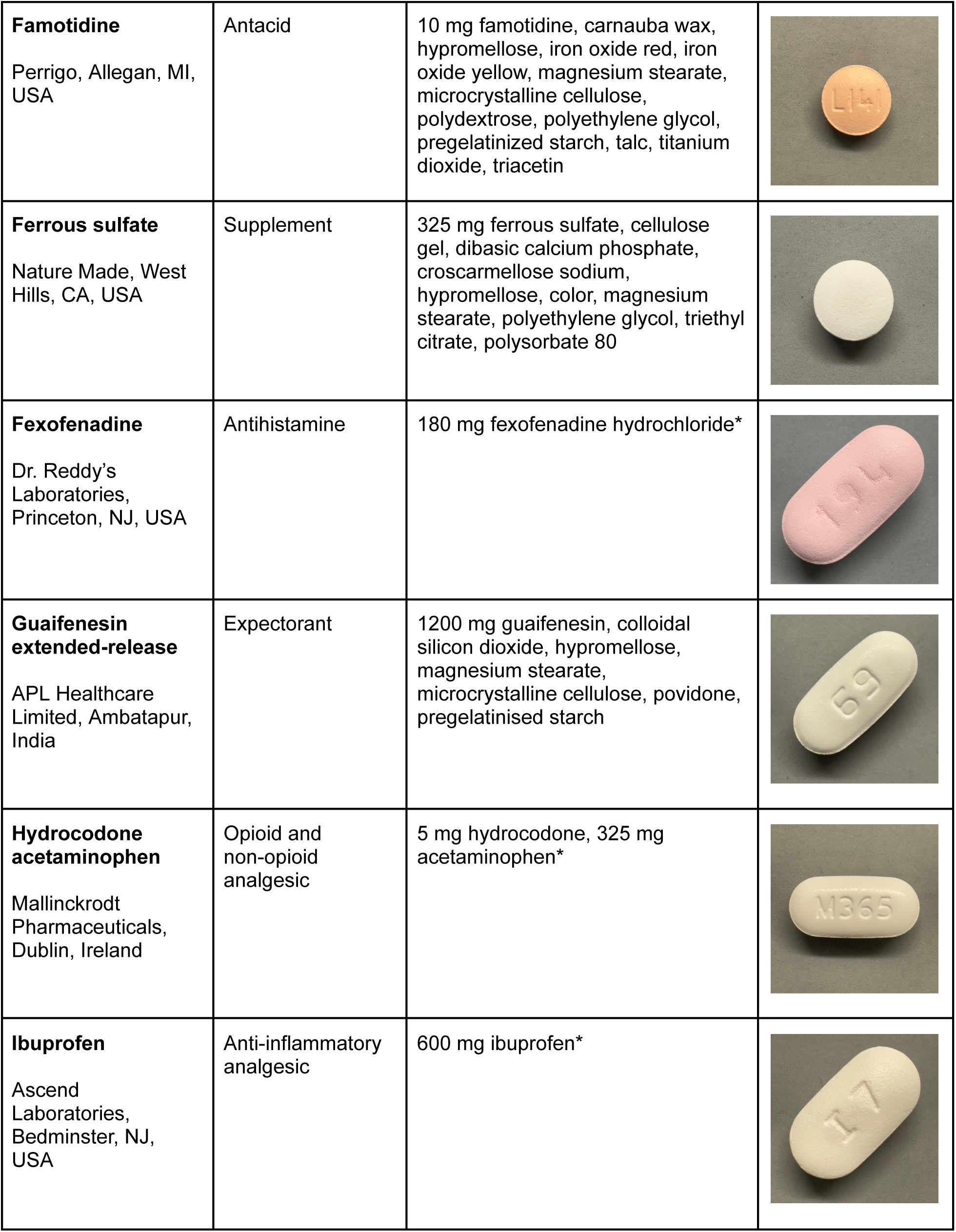
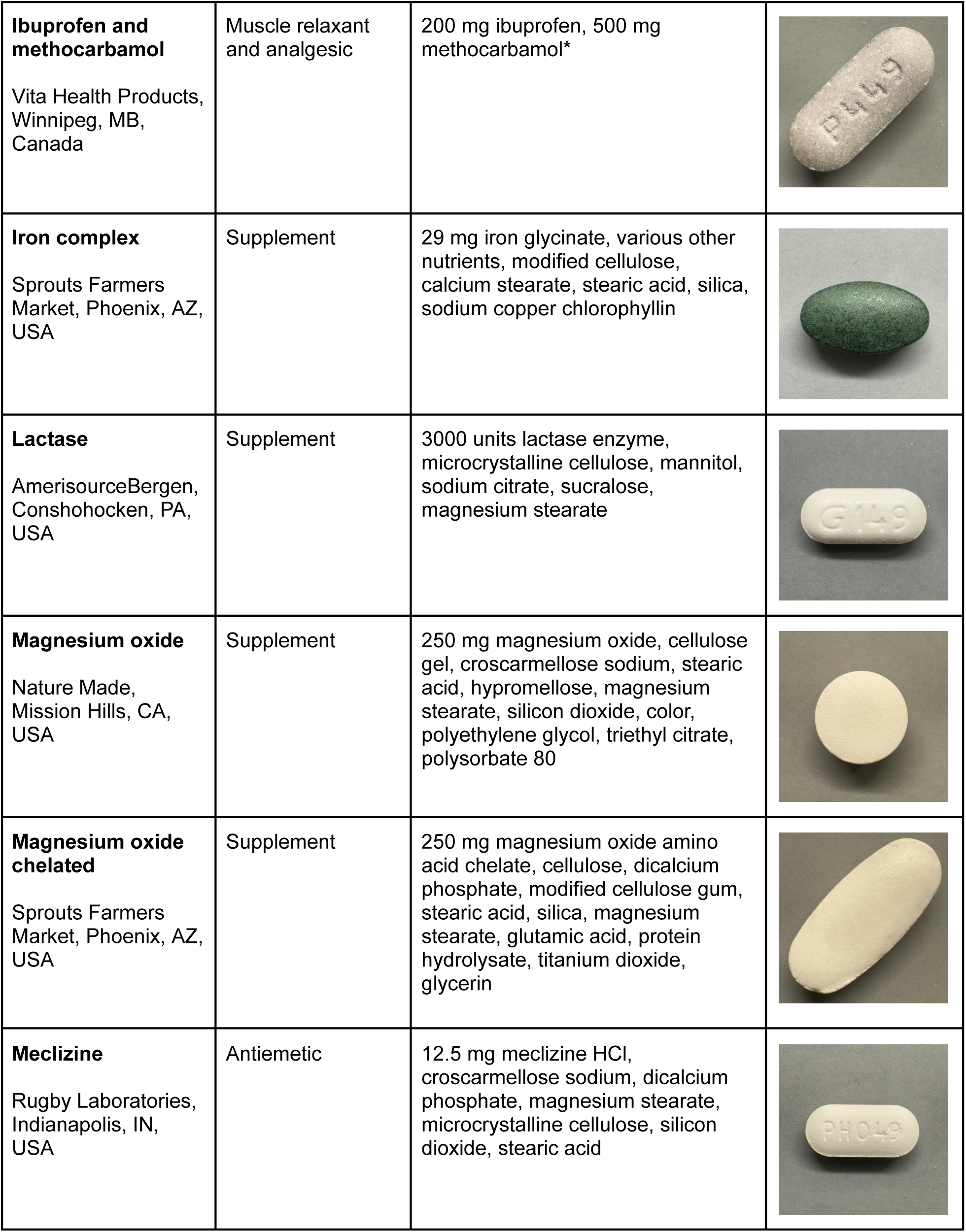
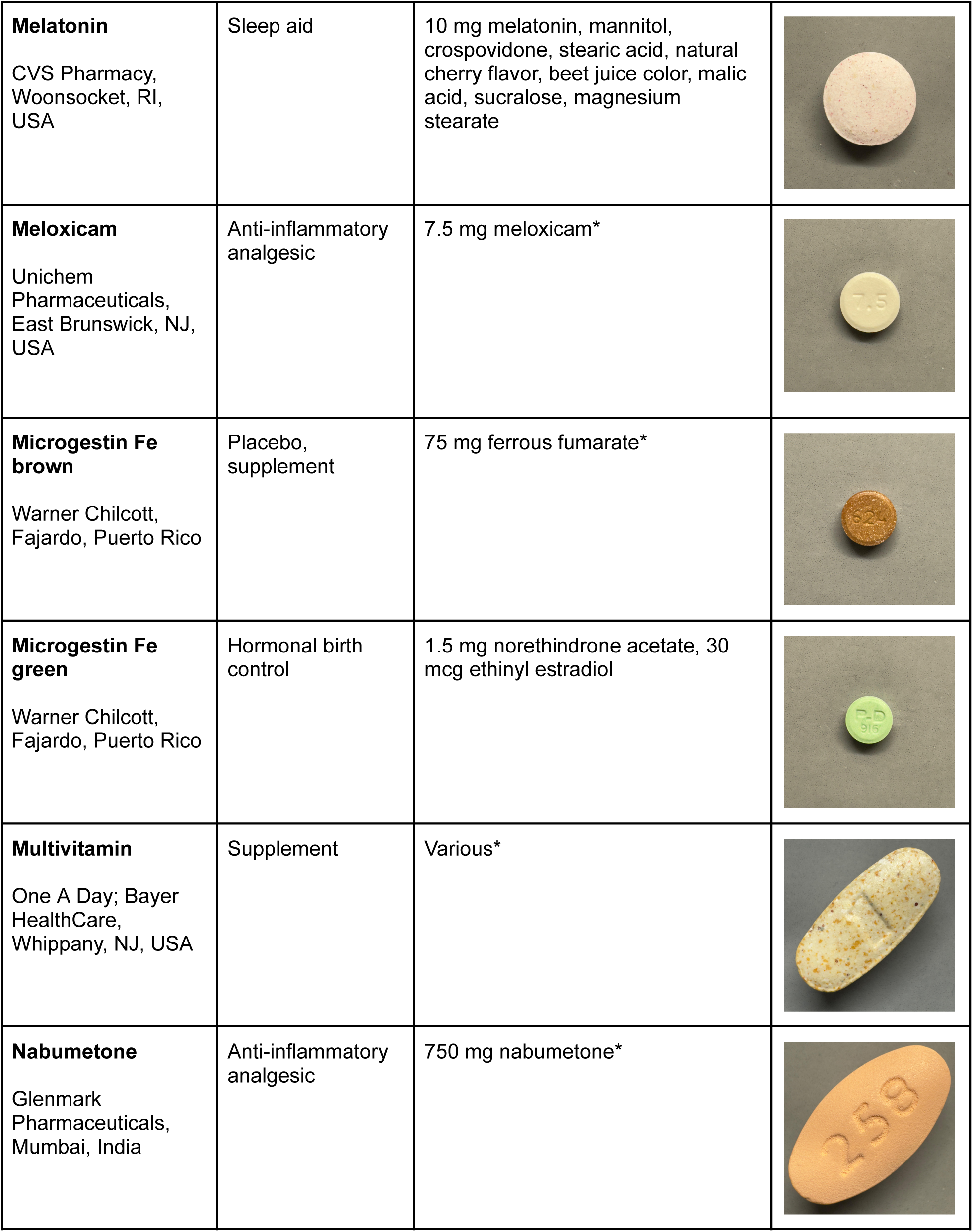
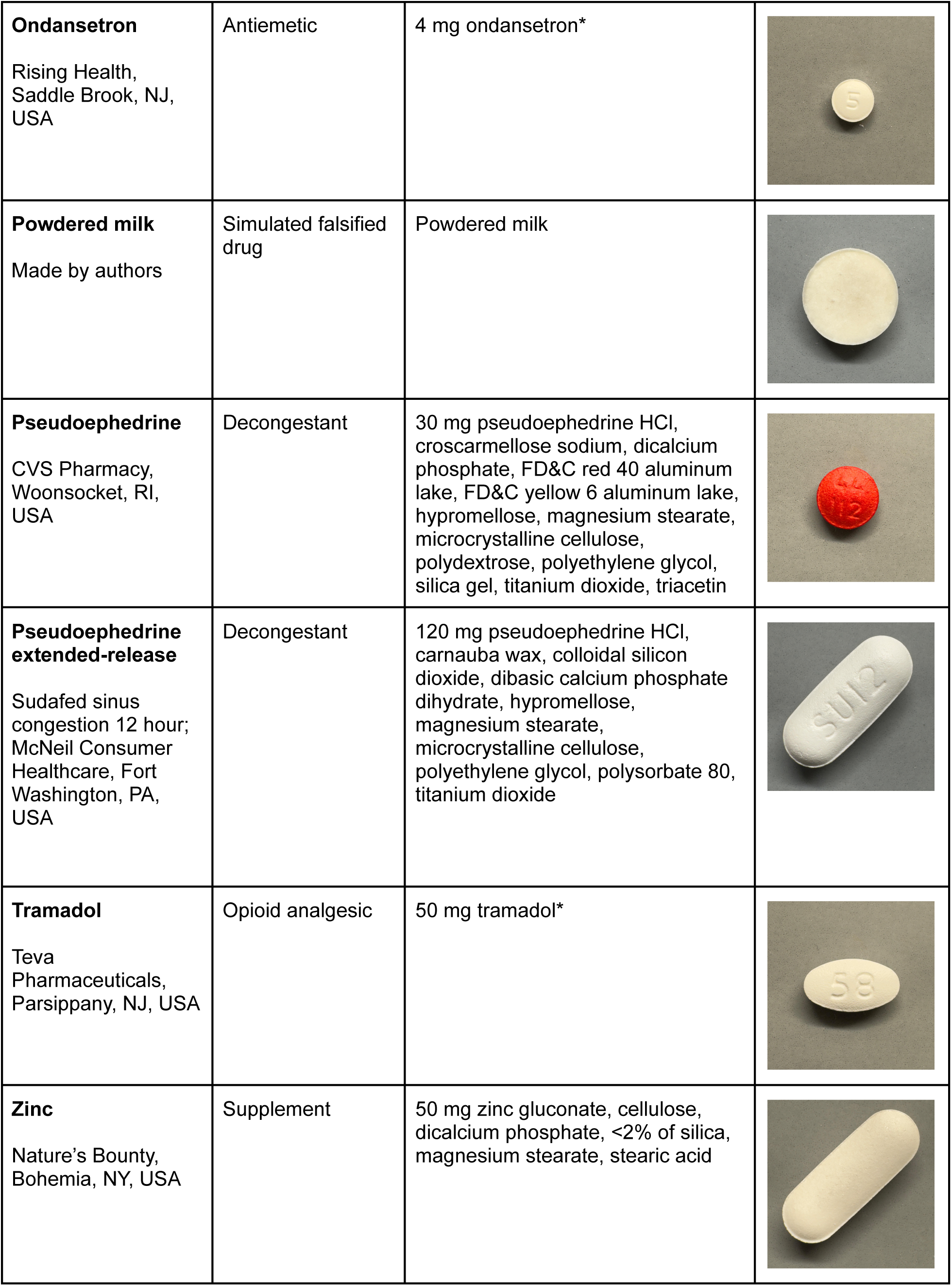
Drug products tested in this study using Disintegration Fingerprinting. * Inactive ingredients not available

| Drug | Purpose | Ingredients | Image |
| --- | --- | --- | --- |
| **Acetaminophen**<br>(Tylenol, USA)<br>McNeil Consumer Healthcare, Fort Washington, PA | Non-opioid analgesic | 500 mg acetaminophen, carnauba wax, castor oil, corn starch, FD&C red no. 40 aluminum lake, hypromellose, magnesium stearate, polyethylene glycol, powdered cellulose, pregelatinized starch, propylene glycol, shellac, sodium starch glycolate, titanium dioxide |  |
| **Acetaminophen**<br>(Tylenol, Canada)<br>McNeil Consumer Healthcare, Markham, ON, Canada | Non-opioid analgesic | 500 mg acetaminophen, cellulose, cornstarch, hypromellose, magnesium stearate, polyethylene glycol, sodium starch glycolate, water |  |
| **Aspirin** (generic)<br>Rite Aid Corporation, Camp Hill, PA | Non-opioid analgesic | 325 mg aspirin, corn starch, hypromellose, polyethylene glycol, propylene glycol |  |
| **Aspirin** (Bayer, USA)<br>Bayer, Leverkusen, Germany | Non-opioid analgesic | 325 mg aspirin, corn starch, hypromellose, powdered cellulose, triacetin |  |
| **Aspirin** (Bayer, Canada)<br>Bayer, Mississauga, ON, Canada | Non-opioid analgesic | 325 mg aspirin, corn starch, hypromellose, powdered cellulose, triacetin |  |
| **B-100 complex**<br>Nature Made, West Hills, CA, USA | Supplement | Various, cellulose gel, hypromellose, stearic acid, magnesium stearate, silicon dioxide, croscarmellose sodium, polyethylene glycol |  |
| **Calcium carbonate**<br>Rugby Laboratories, Livonia, MI, USA | Antacid | 500 mg calcium carbonate, dextrose, flavors, magnesium stearate, maltodextrin, starch, sucralose |  |
| **Cold medicine**<br>(Sudafed PE Pressure Pain Cold); McNeil Consumer Healthcare, Fort Washington, PA, USA | Non-opioid analgesic, decongestant, expectorant, cough suppressant | 325 mg acetaminophen, 10 mg dextromethorphan, 100 mg guaifenesin, 5 mg phenylephrine, carnauba wax, croscarmellose sodium, FD&C yellow 5 aluminum lake, FD&C yellow 6 aluminum lake, hydroxypropyl cellulose, hypromellose, magnesium stearate, microcrystalline cellulose, polyethylene glycol, polysorbate 80, pregelatinized starch, titanium dioxide |  |
| **Doxycycline**<br>Avet Pharma, East Brunswick, NJ, USA | Antibiotic | 100 mg doxycycline* |  |
| **Escitalopram**<br>Zhejiang Huahai Pharmaceutical, Zhejiang, China | Antidepressant | 20 mg escitalopram* |  |
| **Famotidine**<br>Perrigo, Allegan, MI, USA | Antacid | 10 mg famotidine, carnauba wax, hypromellose, iron oxide red, iron oxide yellow, magnesium stearate, microcrystalline cellulose, polydextrose, polyethylene glycol, pregelatinized starch, talc, titanium dioxide, triacetin |  |
| **Ferrous sulfate**<br>Nature Made, West Hills, CA, USA | Supplement | 325 mg ferrous sulfate, cellulose gel, dibasic calcium phosphate, croscarmellose sodium, hypromellose, color, magnesium stearate, polyethylene glycol, triethyl citrate, polysorbate 80 |  |
| **Fexofenadine**<br>Dr. Reddy's Laboratories, Princeton, NJ, USA | Antihistamine | 180 mg fexofenadine hydrochloride* |  |
| **Guaifenesin extended-release**<br>APL Healthcare Limited, Ambatapur, India | Expectorant | 1200 mg guaifenesin, colloidal silicon dioxide, hypromellose, magnesium stearate, microcrystalline cellulose, povidone, pregelatinised starch |  |
| **Hydrocodone acetaminophen**<br>Mallinckrodt Pharmaceuticals, Dublin, Ireland | Opioid and non-opioid analgesic | 5 mg hydrocodone, 325 mg acetaminophen* |  |
| **Ibuprofen**<br>Ascend Laboratories, Bedminster, NJ, USA | Anti-inflammatory analgesic | 600 mg ibuprofen* |  |
| **Ibuprofen and methocarbamol**<br>Vita Health Products,<br>Winnipeg, MB,<br>Canada | Muscle relaxant<br>and analgesic | 200 mg ibuprofen, 500 mg<br>methocarbamol* |  |
| **Iron complex**<br>Sprouts Farmers<br>Market, Phoenix, AZ,<br>USA | Supplement | 29 mg iron glycinate, various other<br>nutrients, modified cellulose,<br>calcium stearate, stearic acid, silica,<br>sodium copper chlorophyllin |  |
| **Lactase**<br>AmerisourceBergen,<br>Conshohocken, PA,<br>USA | Supplement | 3000 units lactase enzyme,<br>microcrystalline cellulose, mannitol,<br>sodium citrate, sucralose,<br>magnesium stearate |  |
| **Magnesium oxide**<br>Nature Made,<br>Mission Hills, CA,<br>USA | Supplement | 250 mg magnesium oxide, cellulose<br>gel, croscarmellose sodium, stearic<br>acid, hypromellose, magnesium<br>stearate, silicon dioxide, color,<br>polyethylene glycol, triethyl citrate,<br>polysorbate 80 |  |
| **Magnesium oxide chelated**<br>Sprouts Farmers<br>Market, Phoenix, AZ,<br>USA | Supplement | 250 mg magnesium oxide amino<br>acid chelate, cellulose, dicalcium<br>phosphate, modified cellulose gum,<br>stearic acid, silica, magnesium<br>stearate, glutamic acid, protein<br>hydrolysate, titanium dioxide,<br>glycerin |  |
| **Meclizine**<br>Rugby Laboratories,<br>Indianapolis, IN,<br>USA | Antiemetic | 12.5 mg meclizine HCl,<br>croscarmellose sodium, dicalcium<br>phosphate, magnesium stearate,<br>microcrystalline cellulose, silicon<br>dioxide, stearic acid |  |
| **Melatonin**<br>CVS Pharmacy,<br>Woonsocket, RI,<br>USA | Sleep aid | 10 mg melatonin, mannitol, crospovidone, stearic acid, natural cherry flavor, beet juice color, malic acid, sucralose, magnesium stearate |  |
| **Meloxicam**<br>Unichem<br>Pharmaceuticals,<br>East Brunswick, NJ,<br>USA | Anti-inflammatory analgesic | 7.5 mg meloxicam* |  |
| **Microgestin Fe brown**<br>Warner Chilcott,<br>Fajardo, Puerto Rico | Placebo, supplement | 75 mg ferrous fumarate* |  |
| **Microgestin Fe green**<br>Warner Chilcott,<br>Fajardo, Puerto Rico | Hormonal birth control | 1.5 mg norethindrone acetate, 30 mcg ethinyl estradiol |  |
| **Multivitamin**<br>One A Day; Bayer<br>HealthCare,<br>Whippany, NJ, USA | Supplement | Various* |  |
| **Nabumetone**<br>Glenmark<br>Pharmaceuticals,<br>Mumbai, India | Anti-inflammatory analgesic | 750 mg nabumetone* |  |
| **Ondansetron**<br>Rising Health,<br>Saddle Brook, NJ,<br>USA | Antiemetic | 4 mg ondansetron* |  |
| **Powdered milk**<br>Made by authors | Simulated falsified drug | Powdered milk |  |
| **Pseudoephedrine**<br>CVS Pharmacy,<br>Woonsocket, RI,<br>USA | Decongestant | 30 mg pseudoephedrine HCl, croscarmellose sodium, dicalcium phosphate, FD&C red 40 aluminum lake, FD&C yellow 6 aluminum lake, hypromellose, magnesium stearate, microcrystalline cellulose, polydextrose, polyethylene glycol, silica gel, titanium dioxide, triacetin |  |
| **Pseudoephedrine extended-release**<br>Sudafed sinus congestion 12 hour;<br>McNeil Consumer Healthcare, Fort Washington, PA, USA | Decongestant | 120 mg pseudoephedrine HCl, carnauba wax, colloidal silicon dioxide, dibasic calcium phosphate dihydrate, hypromellose, magnesium stearate, microcrystalline cellulose, polyethylene glycol, polysorbate 80, titanium dioxide |  |
| **Tramadol**<br>Teva Pharmaceuticals,<br>Parsippany, NJ, USA | Opioid analgesic | 50 mg tramadol* |  |
| **Zinc**<br>Nature's Bounty,<br>Bohemia, NY, USA | Supplement | 50 mg zinc gluconate, cellulose, dicalcium phosphate, <2% of silica, magnesium stearate, stearic acid |  |

To obtain a Disintegration Fingerprint for a pill, the cup is filled with tap water, the stirrer is started, and the pill is placed on the servo platform (Figure 1A). The custom Python software running on a computer connected to the microcontroller records 5 minutes of baseline data from the sensor, then the servo automatically adds the pill to the stirred water (Figure 1B). The software continues to record data from the sensor for an additional 60 minutes. The pill disintegrates into particles, and whenever one of these particles passes near the infrared sensor, the particle reflects some infrared light back into the sensor; this causes a momentary increase in the sensor signal, which results in a peak in the plot of sensor signal vs. time (Figure 1C). For most samples, the number of particles detected in the water varies over the 60-minute run: some samples display more peaks over time (as large particles disintegrate into larger numbers of smaller particles) and some samples have fewer peaks over time (as particles dissolve away and are no longer detected by the sensor). The particular pill analyzed in Figure 1D (an ibuprofen caplet) remains intact for about 1 minute after being added to water, it then disintegrates into a large number of particles by the 3-minute mark, then the number of particles detected slowly drops over the next 20 minutes as the particles dissolve away, but some insoluble particles remain even at the 60-minute mark. By plotting the number of particles detected per minute, we create this pill’s Disintegration Fingerprint shown in Figure 1E. Since the number of particles detected can vary by several orders of magnitude over time for a given pill, we plot DFs using a logarithmic axis for particle count.

## Results and Discussion

### Preliminary validation with insoluble particles

Before using Disintegration Fingerprinting to analyze drug samples, we first wanted to confirm that changes in a DF over time really are caused by changes in the number of particles as a drug disintegrates and dissolves. To verify this, we analyzed particle samples that we know do *not* change over time. For example, Figure 2A shows the DF for a sample of 425-500 µm diameter polyethylene beads in water. Since these beads remain intact and polyethylene is not soluble in water, the number of beads detected should remain constant over time. As expected, the DF for this sample is a flat line. This proves that changes in a DF over time are indeed caused by changes in the number of particles over time, and if these changes in particle count are reproducible for a given drug product, then our technique can be used to “fingerprint” drugs based on their disintegration and dissolution behavior.

**Figure 2:**
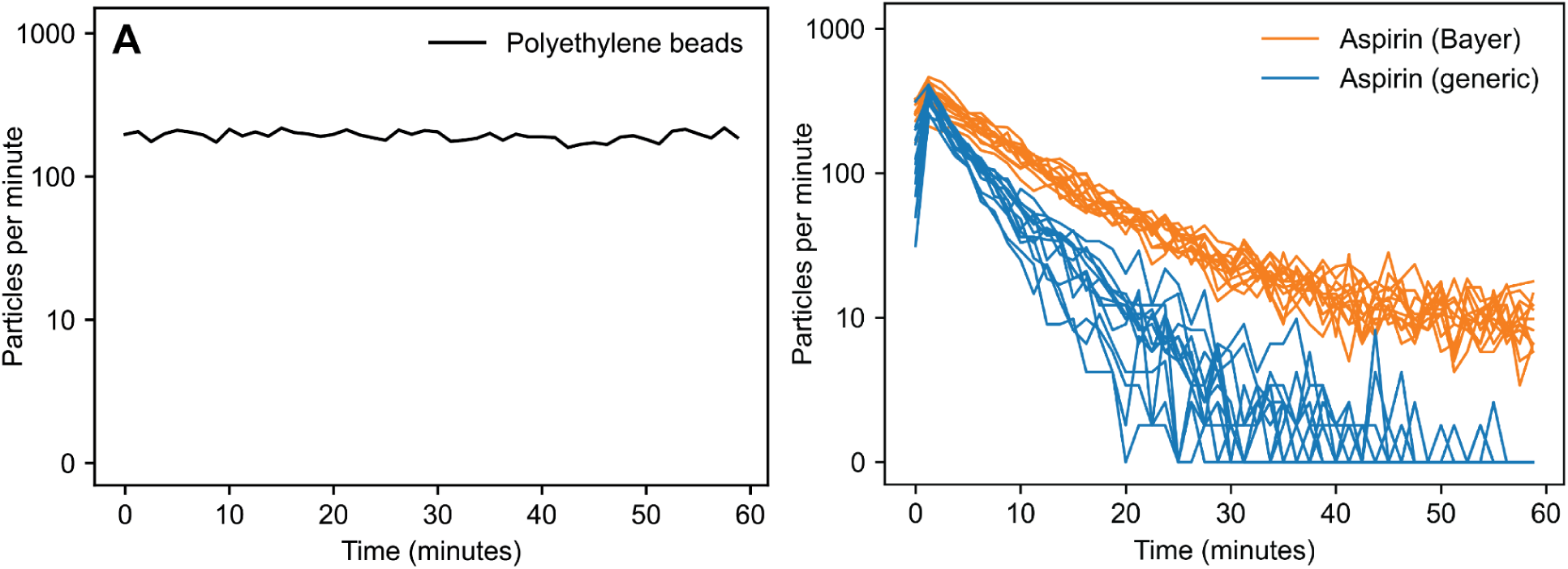
**(A)** The Disintegration Fingerprint (DF) for a sample of polyethylene beads in water is a flat line, indicating that the number of beads is not changing over time (as expected for insoluble polyethylene). In contrast, the DFs for 26 aspirin pills **(B)** exhibit significant change over time as particles disintegrate and dissolve. Even though the 13 pills of name-brand Bayer aspirin (orange traces) and 13 pills of generic aspirin (blue traces) have the same dose of the same active ingredient and similar inactive ingredients, the two products are clearly distinguishable by their DFs.

### Distinguishing name-brand and generic versions of the same product

Next, to explore the ability of Disintegration Fingerprinting to distinguish different products containing the same active ingredient, we next obtained DFs from several aspirin pills (chosen for this testing because aspirin is one of the most widely used drugs worldwide, with tens of billions of aspirin pills consumed every year, and it is a known target for counterfeiters^22^). We analyzed two different aspirin products: a name-brand (Bayer; Whippany, NJ), and a generic (Rite Aid; Philadelphia, PA). Both products have the same amount of active ingredient (325 mg acetylsalicylic acid) and only minor differences in their inactive ingredients (see Table 2 for details), but since they are different products manufactured at different facilities, they should ideally have different Disintegration Fingerprints.

Figure 2B shows Disintegration Fingerprints for 26 aspirin tablets, 13 from a bottle of the name-brand Bayer product (orange traces) and 13 from a bottle of the generic product (blue traces). The different types of tablets are easily distinguished by their DFs: the particle count is consistently higher for the Bayer aspirin with particles still detectable at the 60-minute mark, while the particle count for the generic drops more rapidly and no particles are detectable by the 60-minute mark.

The results in Figure 2B show that these two products can be successfully identified by qualitatively examining their Disintegration Fingerprints. However, a *quantitative* comparison is more valuable because it allows us to rank possible matches by their similarity and calculate the statistical confidence of possible matches. Many different approaches could be used to calculate the similarity of two DFs. In this work, we calculated the sum of the absolute value of the difference between the two DFs. The result, which we refer to as a “difference score,” is relatively small when the two DFs are similar and relatively large when the two DFs are different. Figure 3 shows two examples of difference score calculations. When two Bayer aspirin tablets are compared in Figure 3A, their DFs closely overlap and the resulting difference score (represented by the red area between the two DFs) is relatively low at 861. In contrast, when one Bayer aspirin tablet is compared with one generic aspirin tablet in Figure 3B, the resulting difference score is 2557 (a nearly three-fold increase).

**Figure 3:**
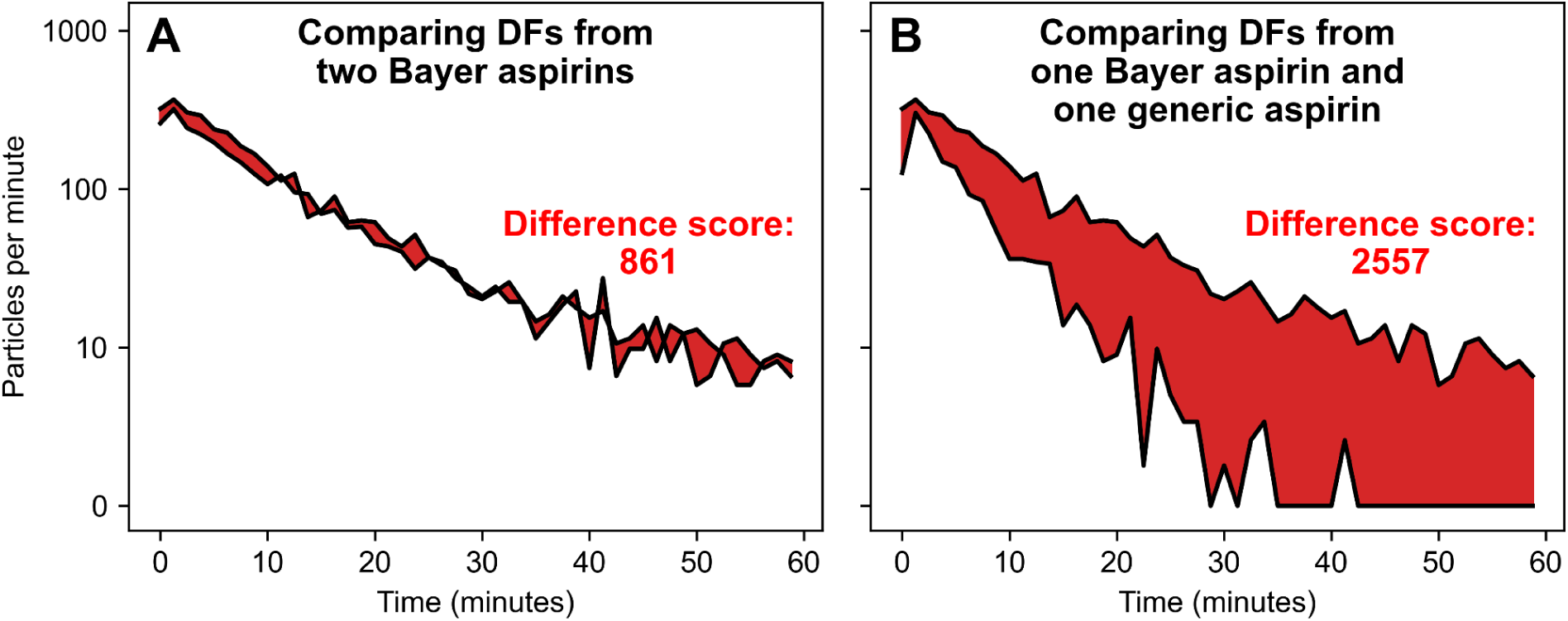
Comparing pairs of Disintegration Fingerprints by calculating “difference scores” represented by the red area between the black DFs. Two Bayer aspirin tablets **(A)** have very similar DFs and consequently result in a lower difference score of 861. In contrast, a Bayer aspirin tablet and a generic aspirin tablet **(B)** have noticeably different DFs and a significantly higher difference score of 2557.

For each of the 26 aspirin DFs in Figure 2B, we calculated the difference score between that DF and each of the other 25 DFs. This resulted in a set of 25 difference scores for each tablet and a total of 325 unique difference scores from all pairwise comparisons of the tablets; these scores are shown in heatmap form in Figure 4. The color of each cell in the heatmap represents the difference score between the two tablets that are in line with the cell’s row and column. Cells with darker blue hues correspond to lower difference scores, and cells with lighter blue hues have higher difference scores. In other words, regions of Figure 4 with darker hues represent pairings of drug products that are more similar to each other according to their DFs, and regions with lighter hues represent products that are more different. The two large dark triangles in Figure 4 confirm that all of the Bayer aspirin tablets were found to be similar to each other (top triangle) and all the generic aspirin tablets were similar to each other (bottom triangle), and the light square region indicates lower similarity between the Bayer and generic tablets. To find the “closest match” for a given tablet in Figure 4, we located the cell in that tablet’s row/column with the lowest difference score (the darkest blue hue) then identified the corresponding product. We found that 100% of these 26 tablets had their closest match to another pill of the same product. Thus, all of these aspirin tablets were successfully identified based on their Disintegration Fingerprints.

**Figure 4:**
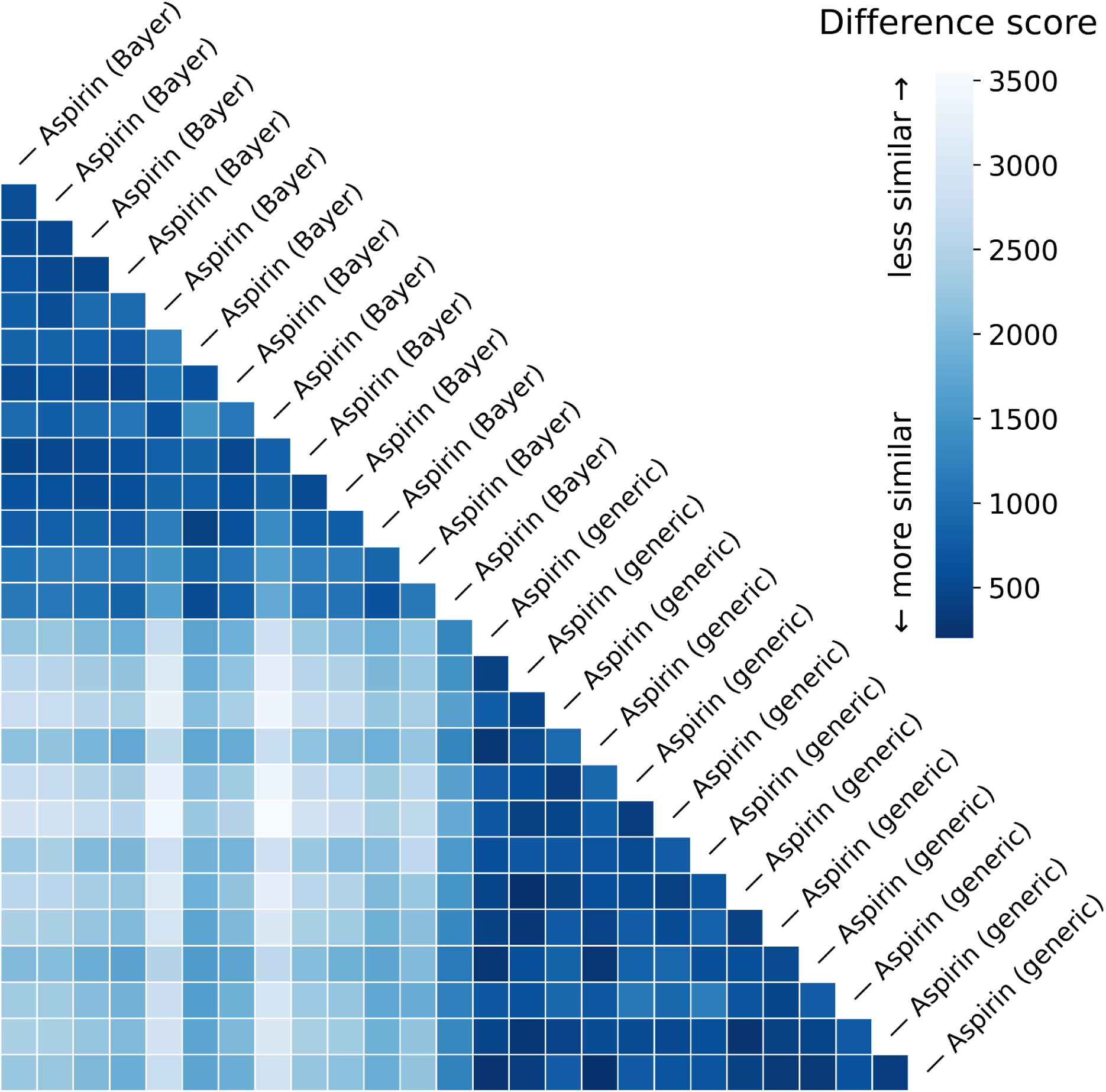
Heatmap of difference scores from all pairwise comparisons of the 26 Disintegration Fingerprints from the 13 Bayer aspirins and 13 generic aspirins shown in Figure 2B. For each pill, the lowest difference scores (dark blue) correspond to other pills of the same product (successful matches), and the highest difference scores (light blue) correspond to pills of a different product.

While our approach was able to correctly identify 100% of the aspirin tablets in our study, Figure 4 does reveal some variability within the difference score values for a given type of comparison: the difference scores corresponding to correct matches are not uniformly dark blue, and the scores corresponding to incorrect matches are not uniformly light blue. To explore this variation, we performed the statistical analysis of the 325 difference scores shown in Figure 5. The results confirm that there is statistically significant variation within the difference scores for each type of comparison. For comparisons between two Bayer aspirin tablets (“Bayer vs. Bayer”), the difference scores range from 404 to 1790 with a median of 820 and an interquartile range of 485; this corresponds to a quartile coefficient of variation of 59%. Similarly, for comparisons between two generic aspirin tablets (“Generic vs. Generic”), the difference scores range from 202 to 1184 with a median of 586 and an interquartile range of 305, which represents a quartile coefficient of variation of 52%. Consequently, the distribution of the “Bayer vs. Bayer” difference scores is significantly different than the “Generic vs. Generic” difference scores (two-sided Mann-Whitney *U* = 4.2 × 10^3^, *p* ≤ 1 × 10^-4^).

**Figure 5:**
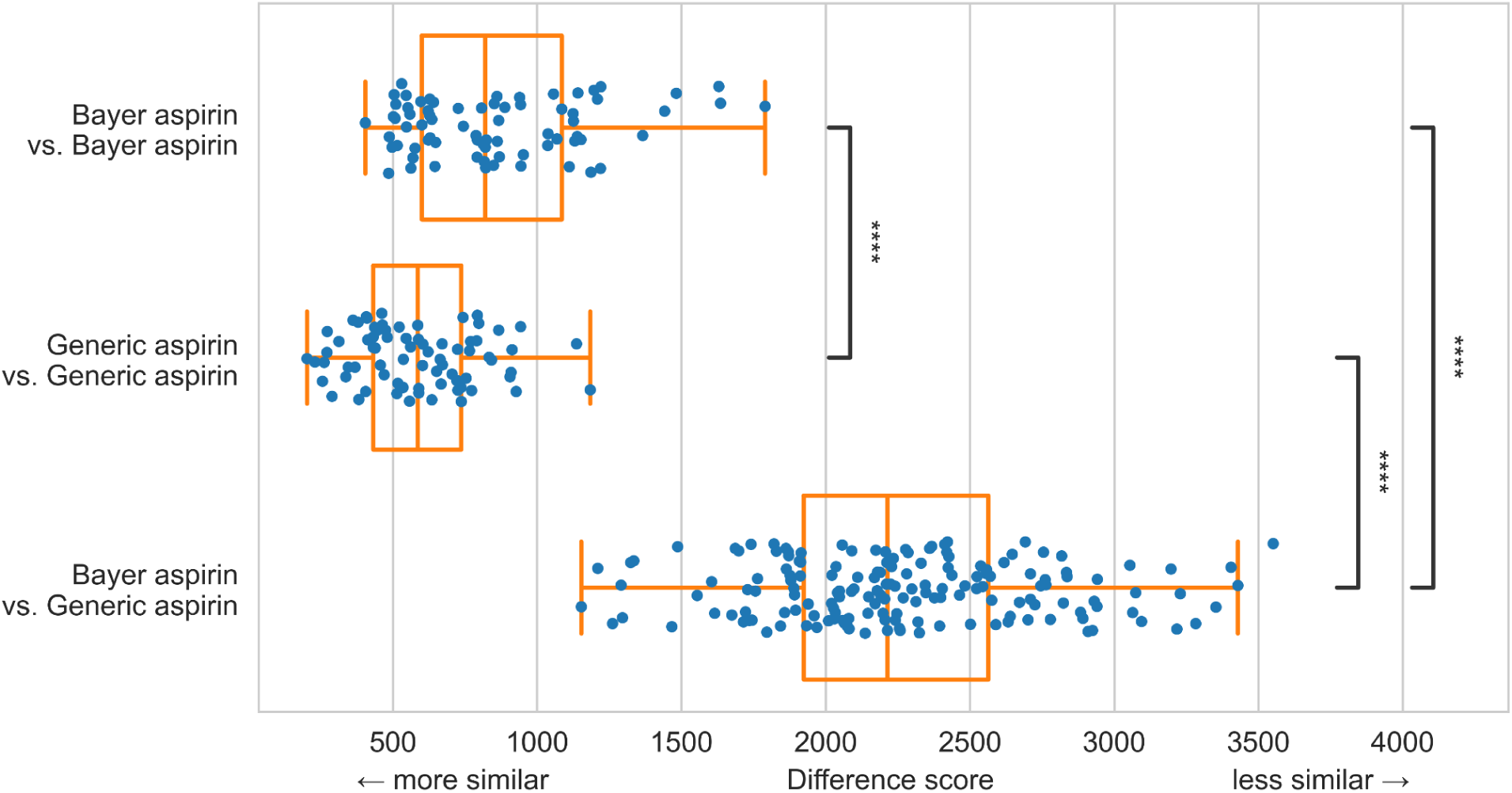
Analysis of the difference scores from the pairwise comparisons of 26 aspirins from Figures 2B and 4. Each blue point corresponds to a difference score from Figure 4, and the orange box-and-whisker plots indicate the median, interquartile range, and upper and lower extremes. While all three distributions of difference scores differed significantly (p ≤ 1 × 10^-4^, details in text), the variability in difference scores among pairs of the same product (Bayer vs. Bayer and Generic vs. Generic) is outweighed by the significantly larger difference scores among pairs of different products (Bayer vs. Generic).

**Figure 6:**
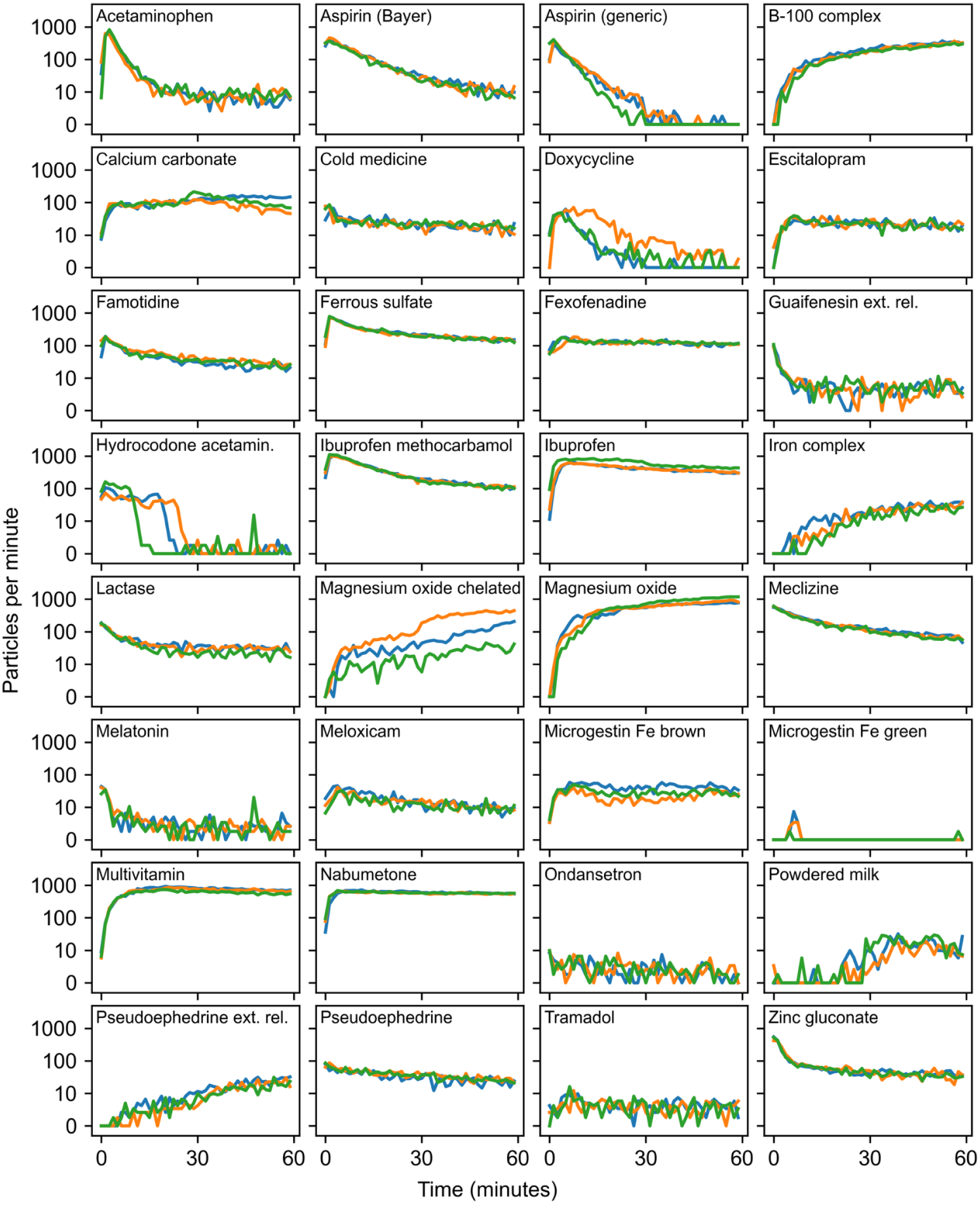
Disintegration Fingerprints for three pills (orange, blue, and green) of each of 32 different drug products. While each product’s DFs are relatively similar, DFs of different products are strikingly different.

However, the comparisons between one Bayer aspirin and one generic aspirin (“Bayer vs. Generic”) generally yield difference scores that are significantly greater than the “Bayer vs. Bayer” and “Generic vs. Generic” scores. The “Bayer vs. Generic” difference scores range from 1153 to 3551 with a median of 2241 (2.7x greater than the “Bayer vs Bayer” median and 3.8x greater than the “Generic vs generic” median), and the interquartile range of the “Bayer vs. Generic” difference scores is 639 (1.3x the “Bayer vs. Bayer” interquartile range and 2.1x the “Generic vs. Generic” interquartile range). Consequently, the distribution of the “Bayer vs. Generic” difference scores is significantly different from both the “Generic vs. Generic” distribution (two-sided Mann-Whitney *U* = 1.0, *p* ≤ 1 × 10^-4^) and the “Bayer vs. Bayer” distribution (two-sided Mann-Whitney *U* = 76, *p* ≤ 1 × 10^-4^). Thus, while there is some variability in difference scores among pairs of the same product, this variability is outweighed by the significantly larger difference scores among pairs of different products.

### Fingerprinting a wider variety of drug products

Next, we wanted to test Disintegration Fingerprinting with a much wider variety of drug products. We used DF to analyze 32 different drug products from a variety of different categories, including antibiotics, opioid and non-opioid analgesics, antidepressants, anti-inflammatories, antiemetics, antihistamines, decongestants, muscle relaxants, expectorants, sleep aids, cold medicines, antacids, hormonal birth control, and dietary supplements. These pills (shown in Table 2) represent an assortment of different sizes and colors, and they include both conventional and extended-release formulations, as well as name-brand and generic manufacturers. Our drug library also included simulated falsified pills that we produced by using a pill press to form powdered milk into tablets.

The results from using DF to analyze our drug library are shown in Figure 6. Each of the 32 plots shows three Disintegration Fingerprints from analyzing three pills of the product shown, for a total of 96 different DFs. Inspecting these plots reveals the striking diversity of the DFs for different drug products, as well as the relative similarity of the three DFs within each drug product. We can also make some qualitative observations about some of the DFs. Some products quickly generate a large number of particles, then the particle count drops as the particles dissolve away; examples of this include acetaminophen, name-brand and generic aspirin, doxycycline, guaifenesin, and hydrocodone acetaminophen. Other products continue to form more particles throughout the entire 60-minute run; these include the B-100 complex, iron complex, chelated and plain magnesium oxide, and pseudoephedrine extended release. The only drug product in our library that generated very few detectable particles (and therefore yielded essentially blank DFs) was the Microgestin Fe green pill; this hormone-containing birth control pill is easily distinguished from the placebo Microgestin

Fe brown pills in the same package based on their DFs. Finally, to facilitate qualitative comparisons between DFs of the different drug products, we calculated the average DF for each drug product and plotted the 32 averaged DFs together on the same axes in Figure 7. The striking diversity of the DFs in Figure 7 supports our claim that different drug products are likely to have different DFs.

**Figure 7:**
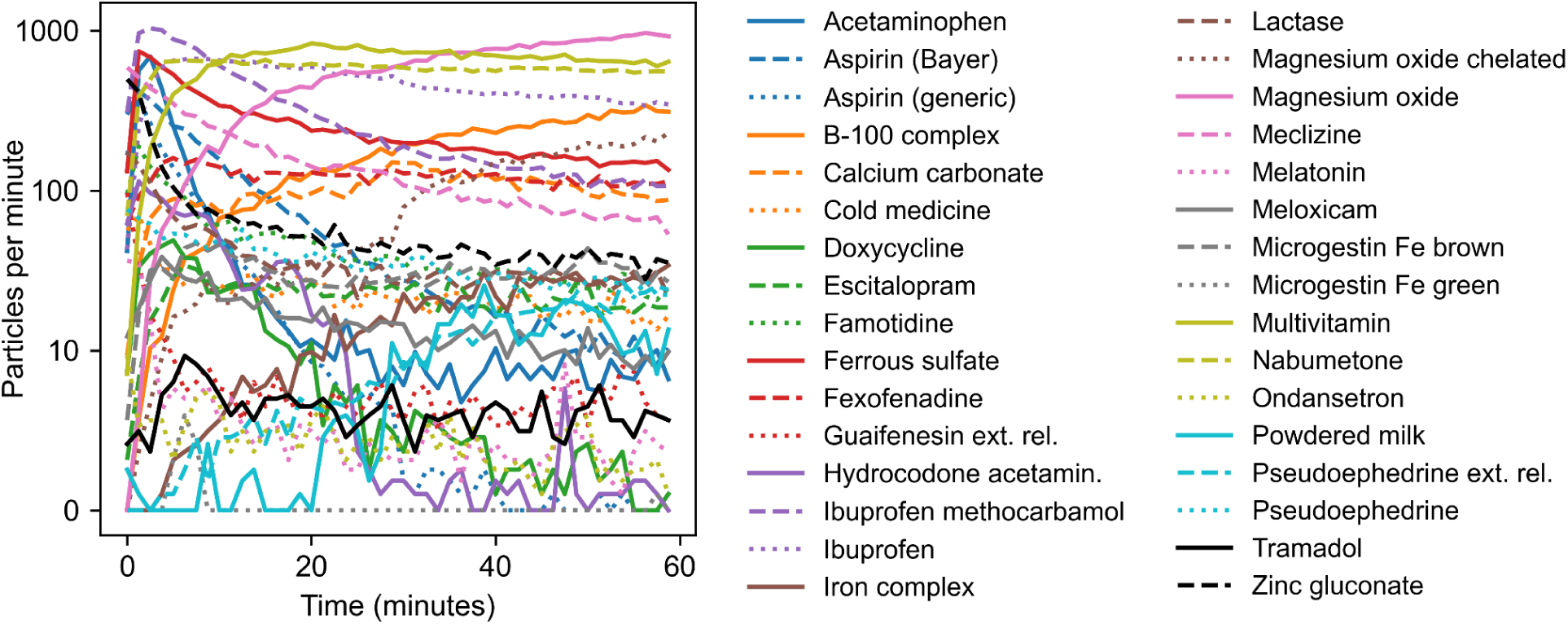
Averaged Disintegration Fingerprints for each of the 32 drug products in Figure 6, showing the large variety of DFs across different drug products.

To quantify the similarity of the products in Figure 6, we again calculated the difference scores between all pairs of DFs. This resulted in a set of 95 difference scores for each pill, as shown on the heatmap in Figure 8. Due to the greater variety of products in this dataset, the difference scores span a much larger range, from a minimum score of 6 (comparing two Microgestin Fe green pills) to a maximum score of 42147 (comparing one Microgestin Fe green pill and one multivitamin pill). Consequently, Figure 8 uses a logarithmic colorbar to help visualize the difference scores that vary by almost four orders of magnitude. As before, dark-blue scores indicate drug pairs with more similar DFs, and light-blue scores indicate pairs with more different DFs. Closer inspection of Figure 8 reveals several sets of three darker-blue difference scores in small “L” shapes that lie along the diagonal line; each of these small dark triangles corresponds to the three comparisons between the three pills of the *same* product. All of the other difference scores in the heatmap correspond to pairings of *different* products. Most of those difference scores are light blue, but some regions of darker blue are evident; these correspond to pairings of different products that nonetheless have relatively similar DFs (in other words, distinguishing those pairs of products is more challenging for our technique).

**Figure 8:**
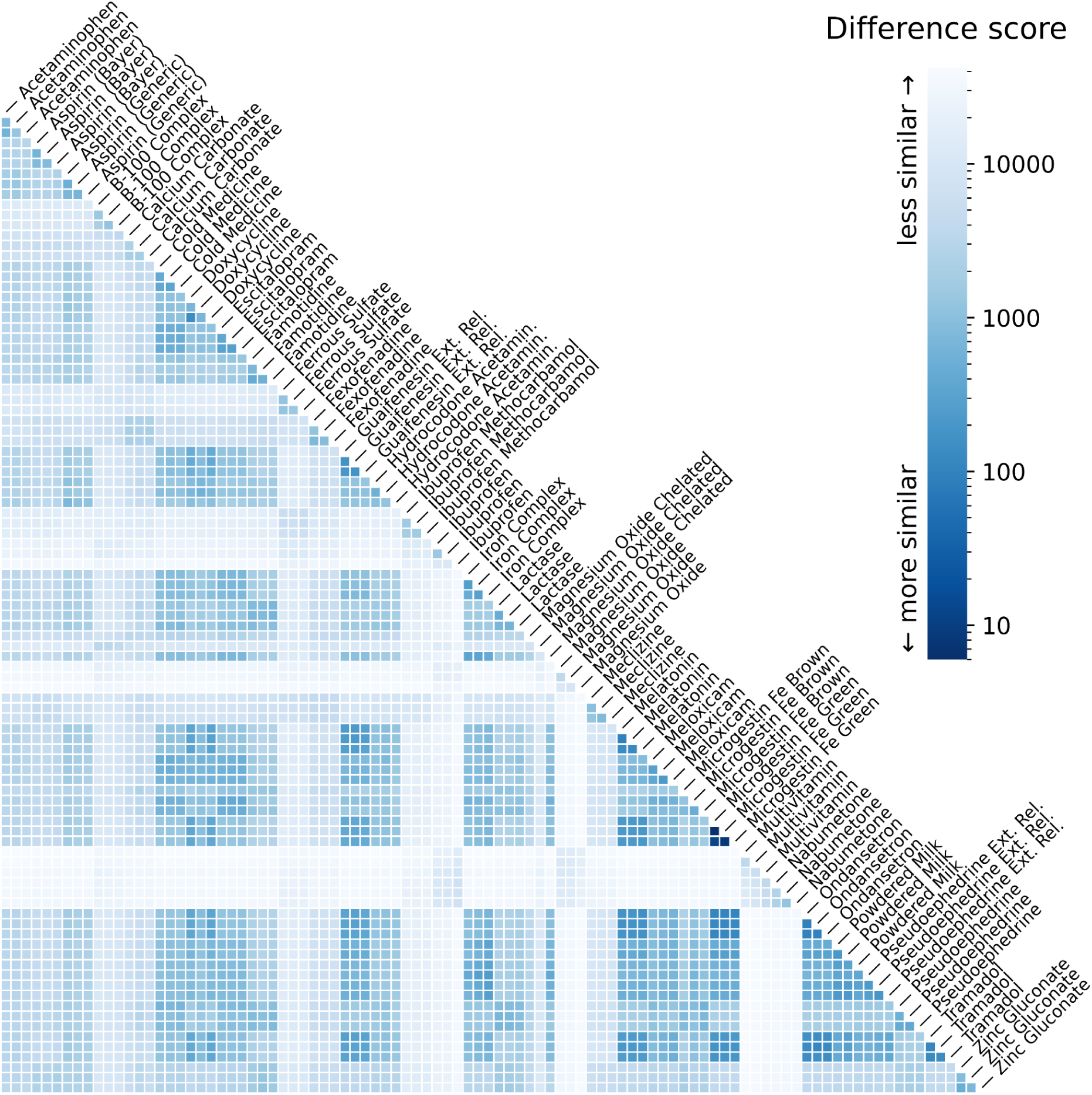
Heatmap of difference scores from all pairwise comparisons of the 96 Disintegration Fingerprints from the 32 drug products shown in Figure 6. Dark blue hues (low difference scores) indicate pairs of pills that have more similar DFs, and light blue hues (high difference scores) represent pairs of pills with less similar DFs. In this heatmap, a logarithmic colorbar is used to span the large range of difference scores.

To find the “closest match” for each pill in Figure 8, we again located the cell in that pill’s row/column with the lowest difference score (the darkest blue hue) and identified the corresponding product. If the closest match was one of the other two pills of the *same* product, this was categorized as a successful identification. If the closest match was one of the 93 pills of a *different* product, this was categorized as an unsuccessful identification. Using this approach, we found that Disintegration Fingerprinting correctly identified 86 of the 96 pills, a success rate of 90%. Of the 10 pills that were misidentified, seven of the pills were the only pills of their product that were misidentified (meaning that the other two pills of that product were correctly identified). The remaining three misidentified pills were all of a single product: magnesium oxide chelated. Examination of the DFs for magnesium oxide chelated in Figure 6 confirms that, for some reason, this product has unusually large variation among its three DFs, which is why comparisons with other drug products had lower difference scores than comparisons with the same product. In summary, for the 32 different drug products in Figure 6, 31 of them (97%) had at least two of the three pills successfully identified using Disintegration Fingerprinting.

Figure 9 plots the distributions of difference scores for each of the 32 drug products in Figure 8. For each drug, each point represents a comparison between one pill of that drug and another pill from the dataset. Comparisons between pills of different products are colored blue, and comparisons between pills of the same product are colored orange. For most of the drug products, the orange points appear on the far-left side of the distribution, confirming that DFs of pills of the same product (matches) generally have the smallest difference scores. Over all 32 products, the median difference score for all matches was 671, compared to a median difference score of 5960 for all mismatches (8.9x greater). The distribution of difference scores for matches was also much narrower, with an interquartile range of 983 compared to 15844 for the mismatches (16.1x greater). As noted earlier, only one product, magnesium oxide chelated, could not be correctly identified using Disintegration Fingerprinting (its orange difference scores from matches are indistinguishable from its blue difference scores from mismatches). For the other 31 products, at least two of the three pills tested were correctly identified using our technique.

**Figure 9:**
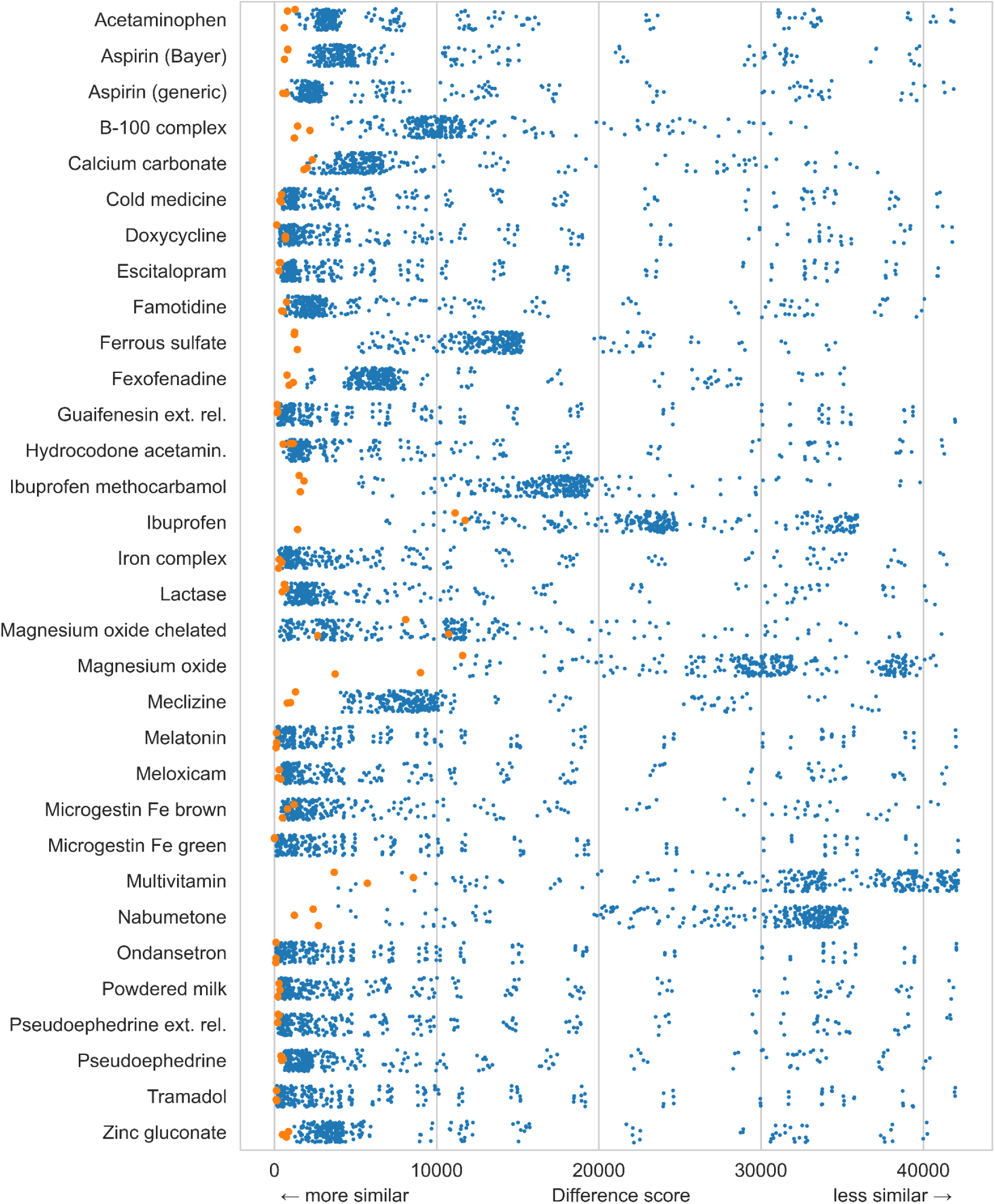
Analysis of the difference scores from all pairwise comparisons of 96 pills from 32 drug products in Figure 6. The difference scores from comparisons between pills of the same product (colored orange) are generally much lower than the scores from comparisons between different products (colored blue); this shows that most of these pills (90%) can be correctly identified by finding the corresponding DF with the smallest difference score.

### Fingerprinting prolonged-release drug formulations

While collecting Disintegration Fingerprints for the 32 different drug products in Figure 6, we observed that four of the products did not fully disintegrate by the end of the 60-minute run. Not surprisingly, all four of these slow-disintegrating products were marketed by the manufacturers as having a “prolonged release,” “time release,” or “extended release” of active ingredients. To see if Disintegration Fingerprinting can offer insights into the behavior of extended-release drug formulations, we obtained 10-hour-long Disintegration Fingerprints for three pills of each of these four drug products. The results in Figure 10 show that the disintegration behavior of these prolonged-release formulations is still changing even hours after being placed in water. One of the B-100 complex pills (blue trace) matched most closely with one of the iron complex pills. The other 11 pills matched correctly with other pills of the same product—a success rate of 92%, similar to what we obtained for the 60-minute runs in Figure 6. Therefore, significantly increasing the duration of the DF does not seem to have a significant effect on the technique’s ability to identify drug products, at least for the products studied here. However, the long-duration DFs in Figure 10 do show that Disintegration Fingerprinting can provide insights into the behavior of controlled-release pharmaceuticals. For example, the guaifenesin pills show two distinct peaks in their DFs, an early release of particles in the first few minutes after the pill is added to water (also visible in Figure 6), and a later secondary release of particles that peaks around the 3-hour mark. Information like this could be useful to pharmaceutical companies’ formulation and quality assurance labs.

**Figure 10:**
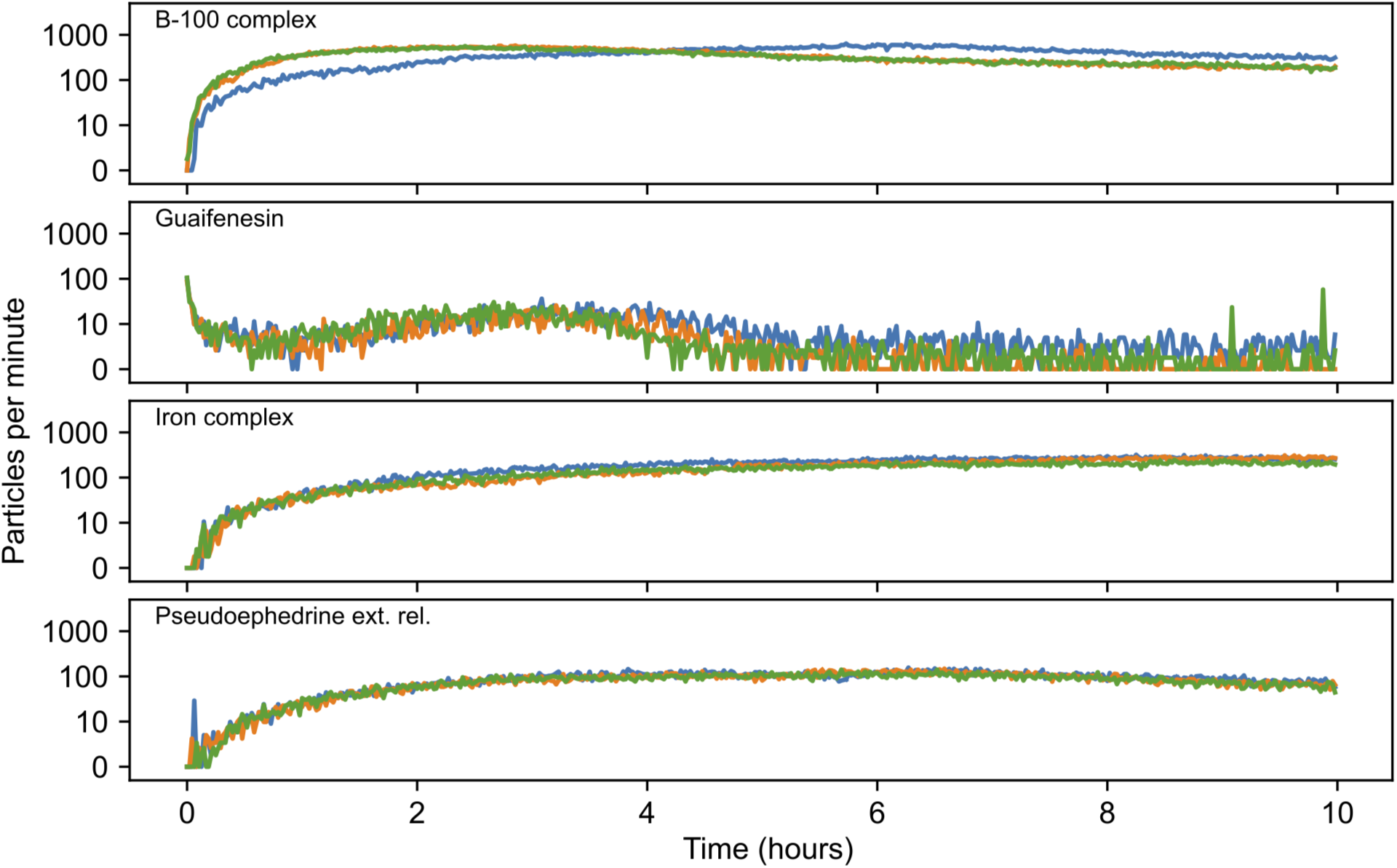
Ten-hour-long Disintegration Fingerprints for three pills (orange, blue, and green) of each of four different extended-release drug products.

### Fingerprinting samples from different manufacturing lots and exposed to extreme temperatures

The Disintegration Fingerprinting technique relies on the assumption that all pills of a given drug product will yield similar DFs. Our results above support this assumption. However, in all of the preceding experiments, when multiple pills of the same product were analyzed, those pills were obtained from a single package. This means that each product’s pills were manufactured at the same time at the same facility, have the same lot number, and have been subjected to the same conditions during distribution and storage. In contrast, different real-world samples of a drug product will likely come from different manufacturing lots, be manufactured at different times (possibly at different facilities), and experience different conditions during distribution and storage. This lot-to-lot variation in a drug product could cause pills from different lots to have different Disintegration Fingerprints, which would make it harder to identify the product using our technique.

To assess the extent to which lot-to-lot variability affects Disintegration Fingerprints, we selected two of the most widely used products from our drug variety, Bayer aspirin (325 mg tablets) and Tylenol Extra Strength acetaminophen (500 mg caplets). We also chose these products because they have fairly similar Disintegration Fingerprints (Figure 6), so even a small amount of batch-to-batch variation in their DFs might adversely affect our ability to distinguish them. We then obtained packages of these products from 33 different manufacturing lots, purchased in eight different states or provinces (British Columbia, California, Colorado, the District of Columbia, Maryland, Minnesota, Tennessee, and Virginia) in two countries (United States and Canada). These products have expiration dates (and, presumably, manufacturing dates) that span almost three years.

Additionally, to explore the effects of different storage conditions on Disintegration Fingerprints, we subjected Tylenol acetaminophen caplets and Bayer aspirin tablets to temperature extremes. Pills from each product were placed in sealed containers and stored at either a high temperature (50 °C) or a low temperature (−20°C) for 35 days. These temperatures are far outside of the recommended storage conditions for room-temperature pharmaceuticals (20-25 °C) and therefore may cause changes in the pills that affect their Disintegration Fingerprints.

The results from analyzing these different manufacturing lots (and different storage temperatures) of Tylenol acetaminophen and Bayer aspirin products are shown in Figure 11. A preliminary visual analysis of the Tylenol DFs shows considerable similarity across the different lots of this product: all of the Tylenol DFs demonstrate an initial spike in particles per minute followed by a rapid drop, with very few particles detected past the 20-minute mark. However, we did notice that two lots, the ones purchased in Canada, seemed to have DFs that were somewhat different from the other DFs (which were from lots purchased in the United States); the Canadian lots had DFs that spike more quickly and drop somewhat more slowly than the US lots. Additionally, all of the Bayer aspirin DFs in Figure 11 looked similar to each other, with a slow decrease in particles per second and a moderate number of particles still detected at the 60-minute mark. The Tylenol and Bayer DFs in Figure 11 can still be distinguished by eye, even though they come from 33 different manufacturing lots, and some were subjected to extreme temperatures.

**Figure 11:**
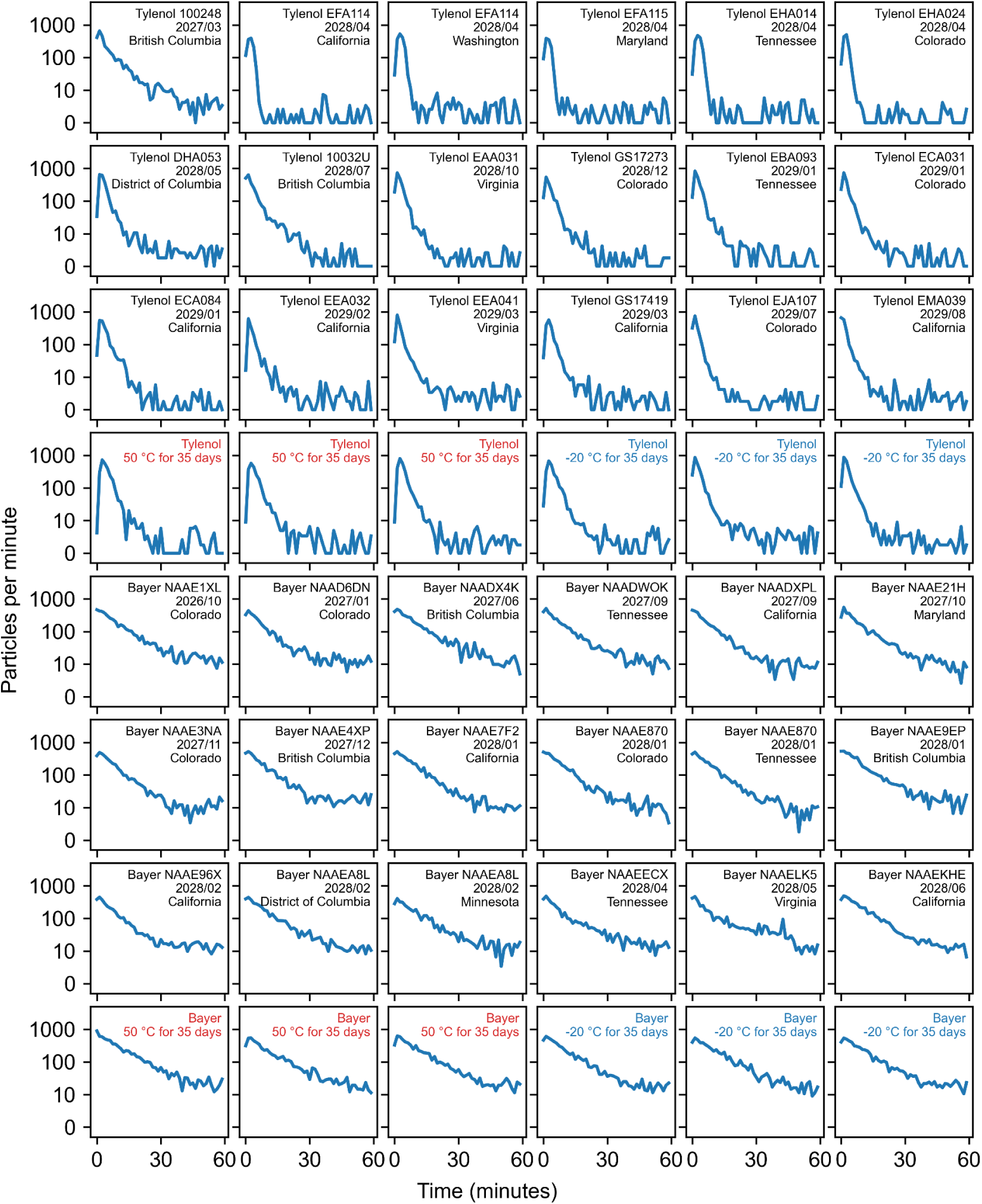
Disintegration Fingerprints from 33 different lots of two drug products, Bayer aspirin (325 mg tablets) and Tylenol Extra Strength acetaminophen (500 mg caplets), purchased in eight different states or provinces in two countries, with manufacturing lot numbers and expiration dates shown. Six pills of each product were also subjected to either high (50 °C) or low (−20°C) temperatures for 35 days. While some lot-to-lot and temperature-induced variation in each product’s DFs may be visible, that variation is outweighed by the overall differences between the two products’ DFs.

For a more quantitative assessment of the similarity of the DFs in Figure 11, we again calculated the difference scores between all pairs of DFs. The resulting heatmap in Figure 12 shows two large dark-blue triangles; these correspond to regions with low difference scores from comparing pairs of Tylenol acetaminophen pills (upper triangle) or pairs of Bayer aspirin pills (lower triangle). Additionally, the light-blue square in Figure 12 corresponds to a region with high difference scores where one Tylenol pill is being compared to one Bayer pill. To find the “closest match” for each pill, we located the cell in that pill’s row/column with the lowest difference score (the darkest blue hue) and identified the corresponding second pill to be the first pill’s closest match. Using this approach, 100% of the pills matched correctly with another pill of the same type. Therefore, at least for these two products, the variation in DFs between the products seems to outweigh the variation between the different manufacturing lots and storage conditions, and Disintegration Fingerprinting is still able to successfully distinguish the products.

**Figure 12:**
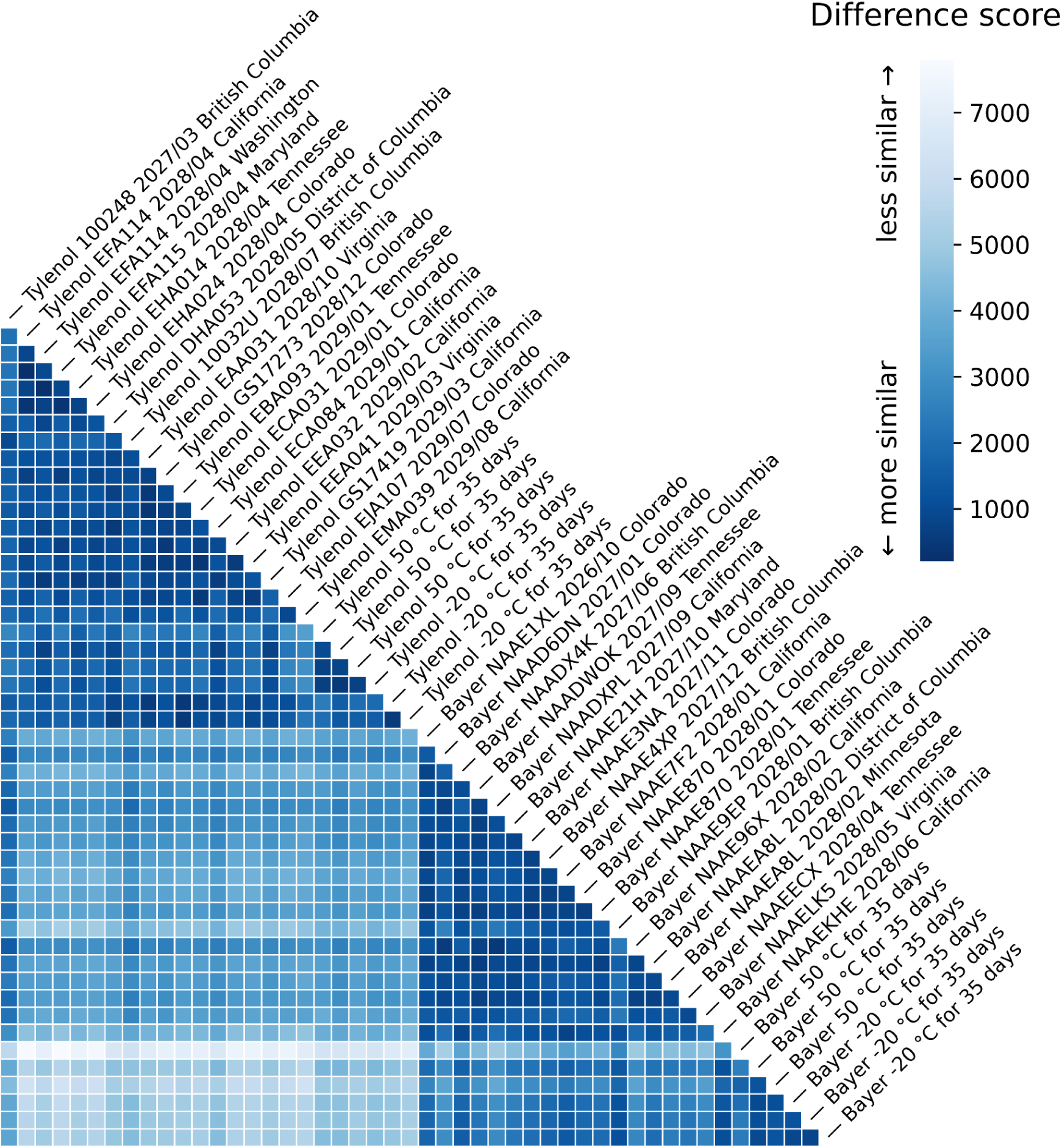
Heatmap of difference scores from all pairwise comparisons of the Disintegration Fingerprints from the 48 pills shown in Figure 11.

Closer inspection of Figure 12 does reveal some potential outliers. For example, the Tylenol acetaminophen caplets from lots 100428 and 10032U seem to have slightly greater similarity to the Aspirin tablets, as evidenced by two darker vertical bars in the first and eighth columns from the left in the square region on Figure 12. Interestingly, these are the Tylenol lots purchased in Canada, so the difference scores corroborate our qualitative observation about the Canadian Tylenol DFs in Figure 11. Additionally, one of the Bayer aspirin tablets that spent 35 days at 50 °C seems to be less similar to both the Tylenol caplets and the other Bayer tablets, as indicated by the lighter horizontal bar on the sixth row from the bottom of Figure 12.

To further explore the trends in difference scores in Figure 12, we plotted the distributions of difference scores for various pairings of different manufacturing lots and storage conditions in Figure 13. In row 1, each point marks the difference score for two Bayer aspirins from the *same bottle* (this is the same distribution as the first row in Figure 5). These same-bottle difference scores have a median value of 820 and an interquartile range of 485. Row 2 shows the corresponding distribution from comparing Bayer aspirins from 16 *different lots*. The difference scores from comparisons across different lots are slightly higher than the same-bottle scores, with a median of 1104 (a 1.3x increase) and an interquartile range of 572. The distributions of difference scores in rows 1 and 2 are statistically significantly different (two-sided Mann-Whitney *U* = 2.2 × 10^3^, *p* ≤ 1 × 10^-4^). Thus, there does appear to be greater variation in Disintegration Fingerprints of aspirins from different lots compared to tablets from a single lot.

**Figure 13:**
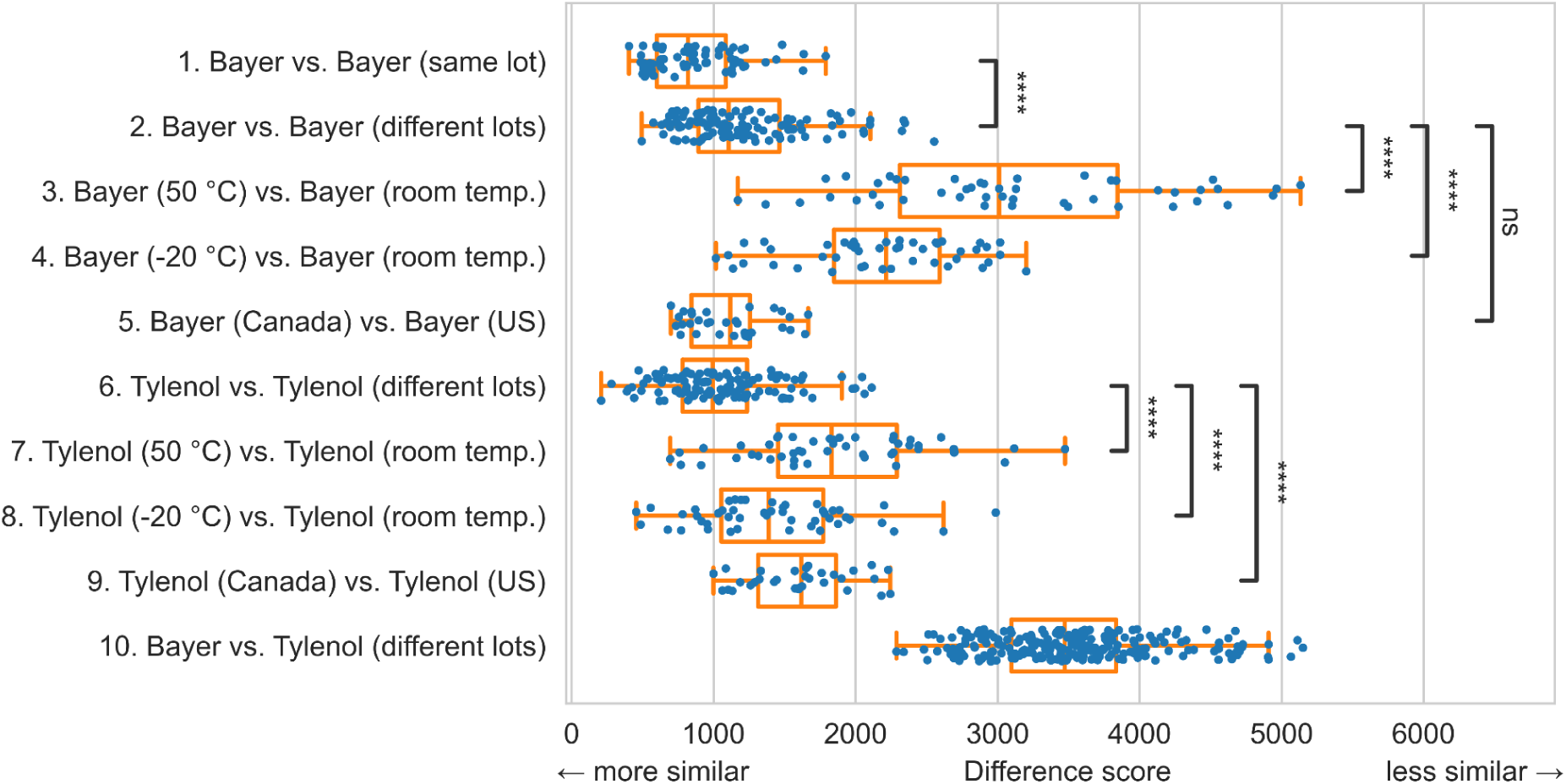
Analysis of the difference scores from the pairwise comparisons of different drug manufacturing lots and storage conditions from Figures 11 and 12. All pairs marked **** had significantly different distributions (p ≤ 1 × 10^-4^, details in text); the pair marked ns was not significantly different (p > 5 × 10^-2^).

However, the Bayer aspirins that were subjected to temperature extremes had much larger difference scores when compared to room-temperature Bayer aspirins. Row 3 in Figure 13 shows the distribution of difference scores when comparing heat-treated Bayer aspirins to room-temperature Bayer aspirins. The difference scores jump to a median value of 3011 (a 2.7x increase over the room-temperature-only difference scores) and the interquartile range increases to 1532 (a 2.9x increase); consequently, the difference score distributions on rows 2 and 3 are significantly different (two-sided Mann-Whitney *U* = 1.8 × 10^2^, *p* ≤ 1 × 10^-4^). Difference scores from comparisons between cold-treated and room-temperature Bayer aspirins (row 4) exhibit a similar increase, with a median value of 2214 (a 2.0x increase over the room-temperature-only difference scores) and an interquartile range of 744 (a 1.3x increase); likewise the difference score distributions on rows 2 and 4 are significantly different (two-sided Mann-Whitney *U* = 5.4 × 10^2^, *p* ≤ 1 × 10^-4^). These results show that exposure to extreme temperatures—especially high temperatures—can have a significant effect on a product’s Disintegration Fingerprint. In the case of aspirin, this temperature sensitivity can be explained by the fact that acetylsalicylic acid readily decomposes via hydrolysis into salicylic acid and acetic acid at elevated temperatures. We weighed Bayer aspirins to determine that each tablet is roughly 82% acetylsalicylic acid by mass; therefore, it is unsurprising that a tablet that is primarily composed of a temperature-sensitive ingredient will behave differently when disintegrating and dissolving in water after exposure to extreme temperatures.

We then examined the distribution of difference scores resulting from comparing Canadian Bayer aspirins to Bayer aspirins from the United States. The results on row 5 of Figure 13 show that the distribution of international difference scores is basically indistinguishable from the distribution of US-only difference scores in row 2. For the “Canada vs. US” difference scores, the median value is 1115 (very similar to the median US-only difference score of 1104) and the interquartile range is 411 (similar to the US-only value of 572). Consequently, the distributions of difference scores on rows 2 and 5 are *not* significantly different (two-sided Mann-Whitney *U* = 2.0 × 10^3^, *p* > 5 × 10^-2^). Therefore, Disintegration Fingerprinting cannot distinguish between Bayer aspirins from these two countries. This is consistent with the fact that Canadian and US Bayer aspirin have the same active ingredient (325 mg acetylsalicylic acid) and the same inactive ingredients (corn starch, hypromellose, powdered cellulose, and triacetin) and suggest that they are identical products (possibly manufactured at the same facility).

Moving on to the distributions of Tylenol acetaminophen difference scores in Figure 13, the median difference score between Tylenols from 17 different manufacturing lots (row 6) was relatively low at 993 (comparable to the median score from comparing Bayer aspirins from the *same* bottle, 820), with an interquartile range of 455. This suggests that the lot-to-lot variability of Tylenol is low, which is consistent with our visual inspection of the Tylenol DFs in Figure 11. Exposing Tylenol to extreme temperatures also increased the difference scores compared to room-temperature Tylenols, although the effect was not as pronounced as it was for the Bayer aspirin. In row 7 of Figure 13, exposure to high temperature caused the median difference score to increase to 1829 (a 1.8x increase over the room-temperature difference scores) with an interquartile range of 836 (a 1.8x increase). Consequently, the distributions of difference scores in rows 6 and 7 are significantly different (two-sided Mann-Whitney *U* = 7.7 × 10^2^, *p* ≤ 1 × 10^-4^). Similarly, in row 8 of Figure 13, exposure to low temperature caused the median difference score to increase to 1388 (a 1.4 increase) with an interquartile range of 720 (a 1.6x increase); thus, the difference score distributions in rows 6 and 8 are significantly different (two-sided Mann-Whitney *U* = 1.7 × 10^3^, *p* ≤ 1 × 10^-4^). Therefore, exposure to extreme temperatures has a measurable effect on the Disintegration Fingerprint of a Tylenol caplet, but the effect is less pronounced than that for Bayer aspirin (perhaps because Tylenol is roughly 83% acetaminophen by mass, and acetaminophen is less temperature sensitive than acetylsalicylic acid).

We then examined the distribution of difference scores from comparing Canadian Tylenol caplets to Tylenol caplets from the United States. The results on row 9 of Figure 13 show that the “Canada vs. US” difference scores are moderately higher than the US-only difference scores: the median “Canada vs. US” difference score was 1616 (a 1.6x increase over the median US-only difference score) with an interquartile range of 548 (a 1.2x increase). Also, the distributions of difference scores in rows 6 and 9 are statistically significantly different (two-sided Mann-Whitney *U* = 5.4 × 10^2^, *p* ≤ 1 × 10^-4^). Thus, Disintegration Fingerprints from Canadian Tylenol are measurably different from DFs from Tylenol from the United States. This is consistent with our earlier observation that the DFs for the Canadian product looked visibly different from the US product in Figure 11. Indeed, the Canadian and US Tylenol products have different inactive ingredients (details in Table 2), so they are different drug products and should have distinguishable DFs.

Having quantified the lot-to-lot and temperature-induced variations in Disintegration Fingerprints from the *same* drug product, we wanted to compare those variations to the differences in Disintegration Fingerprints between *different* drug products. Each point in row 10 of Figure 13 represents the difference score from comparing one Bayer aspirin tablet to one Tylenol acetaminophen caplet (selected from across all 33 different lots of Bayer and Tylenol). The median difference score in “Bayer vs. Tylenol” comparisons, 3472, is higher than any of the other median difference scores in Figure 12. Additionally, as noted above in our analysis of Figure 11, Disintegration Fingerprinting correctly identified 100% of these pills as either Bayer aspirin or Tylenol acetaminophen, despite the fact that they came from 33 different lots, were manufactured over a nearly three year timeframe, and were subjected to extremely high and low temperatures.

Finally, compare the scale of the x-axis on Figure 13 with the same axis on Figure 9 (from our library of 32 different drug products). While the “Bayer vs. Tylenol” comparisons across different lots in Figure 13 have difference scores that range from about 2500 to 5000, the comparisons across the 32 different products in Figure 9 have some difference scores that exceed 40,000. In other words, Bayer and Tylenol are relatively similar products in terms of their Disintegration Fingerprints. Despite this, Disintegration Fingerprinting can still correctly identify 100% of the Bayer and Tylenol pills we tested, regardless of their different lots, different manufacturing dates, and different temperature exposures.

## Conclusions

In this work, we showed that a $42 USD device is capable of correctly identifying 90% of pills in a library of 32 different drug products (and for 97% of the drug products tested, the device successfully identified at least two of the three pills tested). The technique can also successfully distinguish different drug products even if they come from different manufacturing lots, are different ages, and were subjected to extreme temperatures. We conclude by reflecting on some of the strengths, weaknesses, and future directions of the Disintegration Fingerprinting technique.

In addition to its low cost, Disintegration Fingerprinting requires no consumables (reagents or single-use devices); this makes the technique particularly suitable for use in settings without reliable supply chains. Additionally, no laboratory-grade purified water is needed, as we performed our analyses using ordinary municipal tap water. DF can be used to fingerprint *any* solid-dosage drug that disintegrates into particles in liquid. Operating our prototype DF instrument requires no specialized training. Setting up an experiment takes only a few seconds, after which the operation of the instrument is completely automated.

The runtime of our prototype DF instrument, currently 60 minutes per sample, makes our instrument slower than many of the existing techniques shown in Table 1. Obviously, faster is better for many applications, and it may be possible to obtain adequate DFs for some applications in less than 60 minutes. For example, Figure 2B shows that by the 10-minute mark the name-brand and generic aspirin DFs are already clearly distinguishable.

Disintegration Fingerprinting does not provide information about the presence or amount of active ingredients in a drug product. Consequently, DF is best suited for use as a presumptive test for flagging suspect samples for additional confirmatory testing using (much more expensive) tools such as HPLC and MS. Additionally, DF could be very powerful when paired with low-cost techniques that *do* detect active ingredients, such as the aforementioned PADs,^7^ μPADs,^8^ and the GPHF MiniLab.^11^

For 10% of the pills in our test library, Disintegration Fingerprinting did not successfully identify the pill. How could the technique be improved to reduce this error rate? One relatively simple but potentially powerful modification would be to obtain DFs in additional liquids. For example, many drug products have enteric coatings that keep the pill intact in the acidic contents of the stomach but allow the pill to disintegrate in the more alkaline contents of the intestines. By obtaining DFs for a drug product in two different liquids, one with a pH ∼3 and another with pH ∼8, we could verify the presence of an enteric coating based on the resulting DFs. Additionally, since the solubility of many substances is influenced by pH, obtaining DFs in different-pH liquids would make the technique even more sensitive to differences in the chemical compositions of different drug products. Likewise, since solubility is also influenced by temperature, two drug products that have indistinguishable DFs at one temperature might be expected to have different DFs at other temperatures. And since different substances also have different solubilities in different solvents, obtaining DFs in additional *non-aqueous* liquids could further enhance the discrimination ability of the technique (for example, since acetylsalicylic acid is more soluble in ethanol than water, one would expect an authentic aspirin would generate particles that dissolve faster in ethanol than in water).

Another likely source of error in our prototype DF instrument concerns our stirrer. The rate at which a solid-dosage drug disintegrates in water is heavily influenced by how much agitation the water is receiving; for this reason, the various USP dissolution apparatuses^17^ are engineered to apply a precisely-controlled amount of agitation to pills as they disintegrate. In contrast, the magnetic stirrer used in our prototype DF instrument costs just $24 USD and does not provide fine control; we merely adjusted its speed until the vortex extended 1 to 2 cm below the surface of the water. Magnetic stirrers are known to be sources of inconsistencies: researchers found that chemical reactions can have irreproducible results because of variations in the location of a container atop a stirrer.^23^ Therefore, replacing our low-cost magnetic stirrer with a more consistent tool for applying agitation (likely a paddle-based design, like the ones used in some USP dissolution apparatuses^17^) might improve the reproducibility of our technique, albeit probably at greater hardware expense.

The algorithms we used in this preliminary demonstration are simplistic and could be outperformed by more sophisticated methods. There is clearly information present in the raw plot of sensor signal vs. time (Figure 1D) that is absent from the plot of peak count vs. time (Figure 1E). Algorithms that compare pills based on their raw plots of sensor signal vs. time could exploit not only differences in peak counts, but also differences in peak heights, peak widths, baselines, and more. Similarly, the algorithm we used to quantify the similarity between two Disintegration Fingerprints—summing the differences between the curves at each point as shown in Figure 3—is very basic. More sophisticated algorithms that are sensitive to differences in curve shapes (and not just arithmetic differences between peak counts) could outperform the approach shown here.

Our data shows that exposure to extreme temperatures can affect a pill’s Disintegration Fingerprint. Obviously this temperature sensitivity could be detrimental if temperature-induced changes make it difficult to identify a product by its DF (although that was not observed in this study). But this temperature sensitivity could also open up an important new application for Disintegration Fingerprinting. Temperature is said to be the most important environmental factor contributing to a drug’s degradation,^24^ and manufacturers specify recommended storage temperature ranges to protect the therapeutic effectiveness and safety of their products. Studies have shown that exposure to hot conditions significantly decreases the concentrations of the active ingredients in some drug products,^25,26^ and even a brief stint in the trunk of a car on a sunny day could cause damage.^27^ Therefore, there is a need for tools and techniques for identifying drug products that have been exposed to excessive temperatures during storage and distribution. If Disintegration Fingerprinting is consistently sensitive to thermally induced changes in a drug product, then DF could play a role in identifying damaged products before they are consumed by patients. Further inquiry into this topic is ongoing.

Like any drug fingerprinting technique, Disintegration Fingerprinting would benefit from the creation of a large library of DFs from various drug products; a user with a suspect product could obtain its DF and search for a match in the library. Creating this library is beyond the scope of this proof-of-concept study, although we hope that the creation of such a library might be facilitated by the low-cost and open-source nature of our technique’s hardware and software. Even without a library of DFs, a user can always use our technique to analyze two samples and determine if they have similar DFs (which suggests that the samples may be the same product) or different DFs (which is strong evidence that the products are different and the suspect sample should be subjected to additional analysis).

Disintegration Fingerprinting could also play a role in pharmaceutical manufacturers’ quality control programs. For example, a common manufacturing defect involves pharmaceuticals that experience excessive force in the tablet press;^28^ the resulting tablets do not dissolve in the patient’s GI tract as intended, and the patient does not receive the active ingredients. This defect can be detected using USP dissolution apparatuses,^17^ but the complexity and cost of this equipment imposes a practical limit on the number of tablets that can be tested. Disintegration Fingerprinting could serve as a fast and low-cost tool for screening large numbers of tablets for dissolution defects, flagging suspect manufacturing lots for secondary testing using conventional methods.

We are hopeful that Disintegration Fingerprinting can join the ranks of other low-cost and user-friendly tools in the fight against substandard and falsified medicines. Our preliminary results here suggest that the technique has potential, but field testing is a necessary next step. To facilitate widespread evaluation and adoption of DF, all of our CAD files and code are open source and freely available for download.^20^

## Data Availability Statement

All data obtained in this study are freely available for download.^20^

## Supporting Material

Supporting Material is available free of charge:^20^

● CAD files for optional custom components (3D printed fixtures and printed circuit board)
● Arduino code for programming Nano microcontroller clone
● Python code for acquiring, comparing, and plotting Disintegration Fingerprints
● Raw data from experiments shown in Figures 2 through 13

## Notes

The authors declare no competing financial interest.

## Data Availability

All data produced are available online at open.groverlab.org.

https://open.groverlab.org

## Acknowledgments

This work was supported by Award 2131428 from the National Science Foundation Division of Biological Infrastructure (William Grover, PI); Award 2133084 from the National Science Foundation Division of Civil, Mechanical and Manufacturing Innovation

(William Grover, co-PI); and Award 2019362 from the National Science Foundation Division of Computing and Communication Foundations (William Grover, co-PI). Oscar Fajardo was supported by the California Medicine Scholars Program at UC Riverside.

